# Game On or Gone Too Far? Executive Functioning and Habit Learning in Problematic vs. Recreational Gamers

**DOI:** 10.1101/2025.10.08.25334347

**Authors:** Krisztina Berta, Zsuzsanna Viktória Pesthy, Teodóra Vékony, Bence Csaba Farkas, Orsolya Király, Zsolt Demetrovics, Dezső Németh, Bernadette Kun

**Affiliations:** Doctoral School of Psychology, ELTE Eötvös Loránd University, Budapest, Hungary; Institute of Psychology, ELTE Eötvös Loránd University, Budapest, Hungary; Institute of Cognitive Neuroscience and Psychology, HUN-REN Research Centre for Natural Sciences, Budapest, Hungary; Gran Canaria Cognitive Research Center, Department of Education and Psychology, Atlántico Medio University, Las Palmas de Gran Canaria, Spain; Université Paris-Saclay, UVSQ, INSERM, CESP, Villejuif, France; Pôle Pilotage activités et Projets, Direction Générale Adjointe Enfance Famille Santé (DGAEFS), Guyancourt, France; Centre de Recherche en Épidémiologie et en Santé des Populations, INSERM U1018, Université Paris-Saclay, Université Versailles Saint-Quentin, Paris, France; Flinders University Institute for Mental Health and Wellbeing, College of Education, Psychology and Social Work, Flinders University, Bedford Park, SA, Australia; Centre of Excellence in Responsible Gaming, University of Gibraltar, Gibraltar, Gibraltar; Centre de Recherche en Neurosciences de Lyon, INSERM, CNRS, Université Claude Bernard Lyon 1, CRNL U1028 UMR5292, Bron, France; BML-NAP Research Group, Institute of Psychology, Eötvös Loránd University & Institute of Cognitive Neuroscience and Psychology, Research Centre for Natural Sciences, Budapest, Hungary

**Author notes:** senior authors (Dezső Németh, Bernadette Kun). These authors contributed equally: Krisztina Berta and Zsuzsanna Viktória Pesthy. These authors jointly supervised this work: Dezső Németh and Bernadette Kun.

**Keywords:** gaming disorder, recreational gaming, executive functions, statistical learning, behavioral addiction, cognitive profiles, competition theory

## Abstract

Video gaming often sparks controversy, though negative effects are mainly linked to gaming disorder, not gaming itself. Research shows that gaming disorder is associated with reduced executive functioning and greater reliance on habitual processes, while recreational gaming may relate to enhanced cognitive functions. However, comprehensive comparisons of cognitive profiles across gaming behaviors remain scarce. Therefore, we aimed to compare the cognitive functioning of non-gamers (NG), recreational gamers (RG), and gaming disorder risk individuals (GDR). Based on the Internet Gaming Disorder Test scores, 114 participants were classified into NG, RG, or GDR groups. Executive functions were assessed using the Go/No-Go, Counting Span, Digit Span, Card Sorting, 1-back and 2-back tasks. Habit learning was measured with the Alternating Serial Reaction Time task. The GDR group showed reduced working memory, performing worse on the Digit Span task than the NG group, and worse on the Counting Span task than both NG and RG groups. Conversely, the RG group displayed enhanced attention-related performance. No group differences emerged in other executive functions or overall habit learning. Interaction analyses revealed a negative relationship between habit learning and inhibitory control/updating across groups, supporting competition theory, while a positive link between working memory and habit learning in NG and GDR groups suggests possible compensatory mechanisms. Overall, this study underscores that cognitive impairments are linked to gaming disorder rather than gaming itself, while recreational gaming may offer cognitive benefits. These findings provide insights into the distinct cognitive profiles of recreational gamers and those at risk of gaming disorder.

**Highlights:**

- Cognitive profiles differ for gaming disorder vs. recreational gaming.
- Working memory is impaired in individuals at risk of gaming disorder.
- Recreational gaming is linked to enhanced attention-related performance.
- Executive functions and habits compete with each other, regardless of gaming.
- Cognitive systems show both competitive and compensatory links.

## 1. Introduction

While gaming is usually an everyday recreational activity that can enhance daily life and provide social benefits, these positive effects can be overshadowed when gaming turns into addiction, severely impacting quality of life (Brand et al., 2025). In 2019, the World Health Organization (WHO) officially recognized Gaming Disorder as a medical condition in the 11th revision of the International Classification of Diseases (ICD-11), defining it as a persistent inability to control gaming, where it takes precedence over daily activities despite negative consequences. The diagnosis requires significant impairment in personal, family, social, educational, or occupational functioning for at least 12 months (Király et al., 2023; World Health Organization, 2018). Similarly, the DSM-5 acknowledges internet gaming disorder as a condition warranting further research, although it has not yet been officially classified as a clinical disorder (American Psychiatric Association, 2013). Given the growing concern over the potential negative impact of gaming disorder, research has increasingly focused on understanding its underlying cognitive mechanisms. A key question is how the cognitive functions involved in gaming disorder compare to those associated with recreational gaming. To explore this, the present study examines both goal-directed and habitual mechanisms in individuals at risk for Gaming Disorder, recreational gamers, and non-gamers, aiming to provide a comprehensive understanding of these cognitive processes.

Goal-directed and habitual systems play a fundamental role in behavioral adaptation, making them potential key factors in differentiating recreational gaming from gaming disorder (Demetrovics et al., 2022; McKim & Boettiger, 2015; Zhou et al., 2021). Goal-directed processes include core executive functions such as inhibitory control, cognitive flexibility, and working memory (Diamond, 2013; Miyake et al., 2000). These functions are cognitively demanding and serve as the foundation for higher-order abilities like problem-solving (Collins & Koechlin, 2012). In contrast, habitual processes rely more on automatic behaviors and are less influenced by immediate goals and outcomes, making them difficult to override even when they result in negative consequences (Horváth et al., 2022; McKim & Boettiger, 2015; Ostlund & Balleine, 2008). These systems contribute to adaptive behavior by being engaged differently based on the complexity and demands of a given situation (Keramati et al., 2016; Wood et al., 2022). Recreational gaming may enhance this behavioral adaptation, as it often requires players to make quick decisions and manage goal-oriented tasks, thereby engaging executive functions (Kowal et al., 2018; Rupp et al., 2019). In line with this, previous research suggests that gamers often demonstrate comparable or even superior goal-directed executive functioning compared to non-gamers (Colzato et al., 2013; Cudo et al., 2025; Waris et al., 2019). However, these benefits may vary depending on the type of game played (Deleuze et al., 2017; Palaus et al., 2017). One might assume that a stronger reliance on goal-directed processes would correspond to a less active habitual system. However, research suggests that recreational gamers may outperform non-gamers in tasks related to habitual learning (Romano Bergstrom et al., 2012). This implies that gaming may enhance sensitivity to statistical patterns, potentially strengthening the habitual system in ways that complement their goal-directed functioning.

Theoretical models suggest that in addictive disorders, habitual processes often remain intact or even become dominant, while goal-directed functions are frequently impaired (Everitt & Robbins, 2016; Furlong & Corbit, 2018; McKim & Boettiger, 2015). While recreational gaming is often associated with enhanced cognitive functions, these benefits may diminish or even disappear in individuals with gaming disorder, where cognitive impairments are commonly observed. This is reflected in the definition of gaming disorder, where a loss of control over gaming is a key characteristic (World Health Organization, 2018). Inhibitory control is one of the most frequently studied cognitive functions in gaming disorder and has been shown to be moderately impaired, as highlighted by a meta-analysis (Argyriou et al., 2017). In contrast, working memory and cognitive flexibility have been less thoroughly investigated. While some studies report impairments in these functions (Jeong et al., 2016; Jiang et al., 2020; Ngetich et al., 2023; Zhou et al., 2016), others have found no significant differences in task performance (Cudo et al., 2025; Ding et al., 2014; Mukherjee & Banerjee, 2022). This mixed evidence suggests that individuals with gaming disorder may often experience impaired goal-directed processes, potentially leading to a compensatory over-reliance on the habitual system. Supporting this, disrupted thalamocortical communication has been observed in individuals with gaming disorder, which may contribute to an overreliance on habitual systems (Zhou et al., 2021). While Kwon et al. (2024) detected no behavioral imbalance between the two systems, their brain imaging data still indicated altered goal-directed signaling. Using an updated version of their two-step task, a newer study subsequently found both reduced goal-directed (model-based) learning and heightened habitual (model-free) signals in the right inferior frontal gyrus, pointing to habit-biased decision making in GD (Lei et al., 2025).

Comparative studies between gaming disorder and recreational gaming have primarily focused on inhibitory control, with results often showing mixed findings, while research into the broader cognitive profiles of these groups remains limited. For instance, one study has reported that individuals with gaming disorder exhibited weaker inhibitory control and distinct neural activation patterns compared to recreational gamers (Dong et al., 2017). Another study, however, found differences only in response to gaming-related stimuli, emphasizing the role of attentional biases in gaming disorder (Maleki et al., 2024). Meanwhile, some research has found no significant differences between these groups, whether the stimuli were neutral or gaming-related (Jeromin et al., 2016). These inconsistencies, coupled with the limited scope of most studies, highlight the need for more comprehensive investigations into the cognitive profiles of gaming disorder and recreational gaming. Such research is crucial for understanding the underlying mechanisms that distinguish healthy gaming from behaviors indicative of addiction.

To address this gap, the present study broadened the focus beyond isolated cognitive processes by examining both goal-directed and habitual processes, comparing individuals with gaming disorder risk, recreational gamers, and non-gaming individuals. Specifically, we investigated cognitive flexibility, inhibitory control, and working memory, as well as habitual processes, using a paradigm designed to measure implicit learning and habit acquisition (Horváth et al., 2022; Howard & Howard, 1997). Based on prior findings, we hypothesized that the subjects at risk of gaming disorder would exhibit impairments in goal-directed functions compared to both recreational gaming and non-gaming groups. In contrast, the recreational gamers were expected to outperform the non-gaming group on tasks requiring executive functions, reflecting enhanced goal-directed abilities. Regarding habitual processes, we anticipated that the gaming disorder risk group would outperform both the recreational gaming and non-gaming groups, reflecting a stronger reliance on habitual mechanisms commonly associated with addictive disorders. Additionally, we investigated the interaction between the goal-directed and habitual systems across the three groups. According to the competition theory (Pedraza et al., 2024; Poldrack & Packard, 2003), these two systems are generally expected to show a negative relationship, as the dominance of goal-directed processes typically suppresses habitual responses, and vice versa. While previous studies have used various experimental paradigms to explore how the balance between these systems shifts in the context of addiction (Kwon et al., 2024; Lei et al., 2025; Wyckmans et al., 2019; Zhou et al., 2018), the present study offers a novel perspective by focusing on their competition. Although competition between the goal-directed and habitual systems has been documented in healthy controls and in a particular substance use disorder (Pedraza et al., 2024; Virag et al., 2015), this dynamic has not yet been investigated in the context of video gaming behavior, underscoring the novelty of our study.

## 2. Methods

### 2.1 Participants and procedure

Inclusion criteria required participants to be at least 18 years old and to self-report their video gaming habits, which were then used for group classification under the inclusion criteria. Exclusion criteria specified that participants must not have consumed any substances (e.g., alcohol, drugs) on the day of the assessment. One participant was excluded from the analysis due to a likely error in self-reported weekly playtime (140 hours/week), which rendered group assignment unfeasible. Additionally, 21 participants who reported playing video games occasionally (i.e., more than 0 but less than the 14-hour weekly threshold set by our criteria) were excluded from the analyses. The final sample thus consisted of 114 participants (*M*_age_ = 35.39 years; *SD* = 10.11, 64.04% males). Additionally, participants were excluded from specific tasks if their scores fell outside two standard deviations from the mean or if assessment issues occurred (e.g. failure of saving data). For further details, see Supplementary Materials S1. The participants were recruited in two phases as part of a larger study on various behavioral addictions (masked for review). In the first step, control subjects were included in the study who did not play video games. They were recruited from our previous study (masked for review), having agreed at that time to be contacted for future research. In the second phase, we recruited active gamers from the readers/followers of Gamestar, the most popular Hungarian online gaming magazine. During the recruitment process, visitors of Gamestar’s website and Facebook group completed a brief online questionnaire via Qualtrics, which asked about the amount of time they spend playing video games and included the gaming disorder screening test (Ten-Item Internet Gaming Disorder Test; IGDT-10, Király et al., 2017). For group analysis, participants were then categorized into three groups:

- a non-gaming (NG) group (*n* = 41, *M*_age_ = 42.15 years; *SD* = 10.31, 24 females, 17 males; M_gaming time/week_ = 0 hours), who did not play video games at all;
- a recreational gaming (RG) group (*n* = 42, *M*_age_ = 33.83 years; *SD* = 7.54, 9 females, 33 males, M_gaming time/week_ = 29.32 hours), who played video games for at least 14 hours per week but did not meet the IGDT-10 criteria;
- and gaming disorder risk (GDR) group (*n* = 31, *M*_age_ = 28.55 years; *SD* = 7.19, 8 females, 23 males, M_gaming time/week_ = 45.42 hours), who played video games for at least 14 hours per week and scored at or above the literature-defined threshold of 5 on the IGDT-10 questionnaire (Király et al., 2017).

In our sample, the majority of participants (71.05%, *n* = 81) resided in the capital city, while 15.79% (*n* = 18) lived in another city or town, 9.65% (*n* = 11) were from a village or hamlet, and 2.63% (*n* = 3) were based in a county seat city. Regarding educational background, 62.28% (*n* = 71) had obtained a college or university degree, 4.38% (*n* = 5) had a doctoral degree, 28.07% (*n* = 32) had completed high school, and 3.51% (*n* = 4) had finished vocational training without a high school diploma. One participant (0.88%) had completed lower secondary education. Data on both residence and education were missing for one individual. Further demographic details by groups are presented in Table 1.

**Table 1.** Sociodemographic characteristics of the sample: residence and education data across NG, RG, and GDR groups.

| | Total sample<br>($n = 114$) | NG group<br>($n = 41$) | RG group<br>($n = 42$) | GDR group<br>($n = 31$) |
| --- | --- | --- | --- | --- |
| Education<br>n (%) |  |  |  |  |
| Lower<br>secondary<br>education | 1 (0.88) | 0 (0) | 0 (0) | 1 (3.23) |
| Vocational<br>training<br>without<br>high school<br>diploma | 4 (3.51) | 0 (0) | 3 (7.14) | 1 (3.23) |
| High school | 32 (28.07) | 4 (9.76) | 13 (30.95) | 15 (48.39) |
| College or<br>university<br>degree | 71 (62.28) | 34 (82.93) | 25 (59.53) | 12 (38.71) |
| Doctoral<br>degree | 5 (4.38) | 3 (7.32) | 1 (2.38) | 1 (3.23) |
| Missing | 1 (0.88) | 0 (0) | 0 (0) | 1 (3.23) |
| Residence |  |  |  |  |
| n (%) |  |  |  |  |
| Village or hamlet | 11 (9.65) | 3 (7.32) | 3 (7.14) | 5 (16.13) |
| Another city or town | 18 (15.79) | 8 (19.51%) | 5 (11.91) | 5 (16.13) |
| County seat city | 3 (2.63) | 1 (2.44) | 1 (2.38) | 1 (3.23) |
| Capital city | 81 (71.05) | 29 (70.73) | 33 (78.57) | 19 (61.28) |
| Missing | 1 (0.74) | 0 (0) | 0 (0) | 1 (3.23) |

*Note.* NG = Non-Gamer; RG = Recreational Gamer; GDR = Gaming Disorder Risk

Participants attended a 1.5- to 2-hour face-to-face session, which began with a briefing followed by the signing of an informed consent form. The session comprised neuropsychological tasks (detailed in the Measures section), sociodemographic questions, and a self-report questionnaire, which participants completed via the Qualtrics platform. To assess video game usage, participants reported their average gaming time on weekdays and weekends, specifying the number of hours played on each. Total weekly playtime was then calculated as: (5×weekday hours)+(2×weekend hours). As compensation, participants received gift vouchers worth €35.

The data were processed anonymously, and the tasks were neither physically nor mentally demanding for participants. The study complied with established ethical research guidelines and was approved by the Institutional Research Ethics Committee (registration number 2020/401). All procedures followed the ethical principles set forth in the Declaration of Helsinki.

### 2.2 Measures

The computational tasks in this study were developed using the JavaScript library jsPsych (de Leeuw, 2015; Vékony, 2021/2021, 2021, 2022, 2021/2022). Participants performed these tasks on a laptop using a QWERTZ keyboard, where non-essential keys were removed to prevent accidental key presses.

#### 2.2.1 Go/No-Go

We measured inhibitory control using a Go/No-Go task where participants viewed a 2 × 2 square grid, each cell containing a blue star (Bezdjian et al., 2009). A letter, either P or R, would randomly replace one of the stars. Participants were instructed to press the spacebar when the letter P appeared (Go trials) and to withhold their response when the letter R appeared (No-Go trials). The task begins with a 20-trial practice session where participants receive feedback on their responses. Following this, the main task starts without feedback, consisting of 160 trials—128 Go-trials and 32 No-Go trials—maintaining an 80:20 ratio. Each trial is displayed for 500 ms (or until a response is made), with a 1500 ms interval between trials. Midway through the task, the instructions were reversed: press the spacebar for R and refrain from pressing for P. The instructions are followed by a 20-trial practice session with feedback. The second part then begins, consisting of another 160 trials with an 80:20 ratio of Go to No-Go trials. Performance was measured by three metrics: correct responses to Go trials (hits), incorrect responses to No-Go trials (false alarms), and d-prime score, which calculates the standardized value of the difference between hits and false alarms (higher values indicating better inhibitory control), as follows:

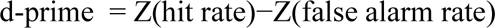

#### 2.2.2 Digit Span task

To assess phonological working memory capacity, we employed the Digit Span Task (DSPAN; Jacobs, 1887; Racsmány et al., 2006). In this task, the examiner verbally presents a sequence of digits, and the participant is required to memorize and then accurately recall the sequence in the same order it was presented. The task starts with recalling a sequence of three digits, with four different sets of these sequences. If the participant correctly recalls at least three of these, they advance to four-digit sequences, and so on. The digit span score is determined by the longest sequence length the participant successfully completes.

#### 2.2.3 Counting Span task

The Counting Span Task (CSPAN; Case et al., 1982) is used to evaluate complex working memory. In this task, participants see various geometric shapes—yellow and blue squares and circles—on the screen. They are required to count the blue circles aloud and remember the total count. This process is repeated across several images. After all images are shown, a question mark appears, prompting the participant to recall and report the sequence of numbers corresponding to the counts they previously repeated, in the correct order. The exercise begins with three practice sets, each containing two trials of geometric shapes. The task then starts with a sequence of two trials, and after a correct response, it progresses to longer sequences, adding one trial at a time, up to a maximum of six trials. This sequence is repeated three times in total. In each round, we recorded the length (i.e., the number of digits that had to be memorized) of the longest sequence that the participant reproduced correctly. The counting-span score was calculated as the average of these three longest-sequence lengths.

#### 2.2.4 N-back tasks

The N-back tasks (Kirchner, 1958) are designed to assess complex working memory, with a particular focus on the updating function of executive processes. In this task, participants view a sequence of letters presented one at a time at the center of the screen. They must determine if the current letter matches the one shown one position earlier (1-back) or two positions earlier (2-back). For matching letters, they press the "J" key; for non-matching letters, they press the "F" key. We set the target ratio at 20%, with 20 targets and 80 non-targets per level. The 1-back and 2-back tasks were administered consecutively, each beginning with a 10-trial practice session that included feedback. Afterward, 100 letters were presented, each displayed for 500 ms or until a response was made, with a 1500 ms interval between them. For analysis, we assessed hit, false alarm, their reaction times, and the d-prime score to evaluate performance.

#### 2.2.5 Card Sorting task

Cognitive flexibility was assessed using the Card Sorting task (CST; Berg, 1948; Fox et al., 2013). In this task, participants are shown four cards in the center of the screen, each displaying geometric shapes that vary by number, color, and shape. A card appears below these, and the participant must match it to one of the top cards based on a specific rule (color, number, or shape). The rule is not given in advance and changes periodically; participants must deduce the rule from feedback indicating whether their match was correct. Participants categorize a total of 64 cards, with the rule changing every 10 cards. Perseverative errors, where participants continue using an outdated rule, indicate challenges in cognitive flexibility. The fewer such errors, the better their cognitive flexibility is considered.

#### 2.2.6 Alternating Serial Reaction Time task

To measure habit learning, we used the Alternating Serial Reaction Time task (ASRT; Howard & Howard, 1997; Song et al., 2007), which implicitly assesses the learning of sequential patterns, essential for habit formation (Horváth et al., 2022). Participants interacted with a screen displaying four circles, each linked to a specific key (‘S’, ‘F’, ‘J’, ‘L’). Images of dog heads would appear as target stimuli in the circles, and participants were instructed to press the corresponding key as quickly and accurately as possible.

The stimuli followed a hidden alternating pattern: every first stimulus was randomly selected, while every second stimulus followed a set sequence. Each sequence consisted of eight elements, such as r 2 r 4 r 1 r 3, where numbers represent pattern-based stimuli, and r denotes random stimuli that could appear in any of the four positions. From this structured sequence, high- and low-probability triplets can be identified. A high-probability triplet (e.g., 2 r 4 or 4 r 1) occurs more frequently because it can be formed both when the triplet begins with a pattern-based stimulus and when it begins with a random one, together accounting for 62.5% of all triplet occurrences. In contrast, a low-probability triplet (e.g., 4 r 2 or 1 r 4) can only be formed when both the first and last elements are random, resulting in a lower frequency of 37.5%. As a result of this distribution, participants are typically faster and more accurate in responding to high-probability triplets, which serves as a behavioral measure of implicit statistical learning (see further details in Supplementary Materials S2; Fig. 1).

**Fig. 1.**
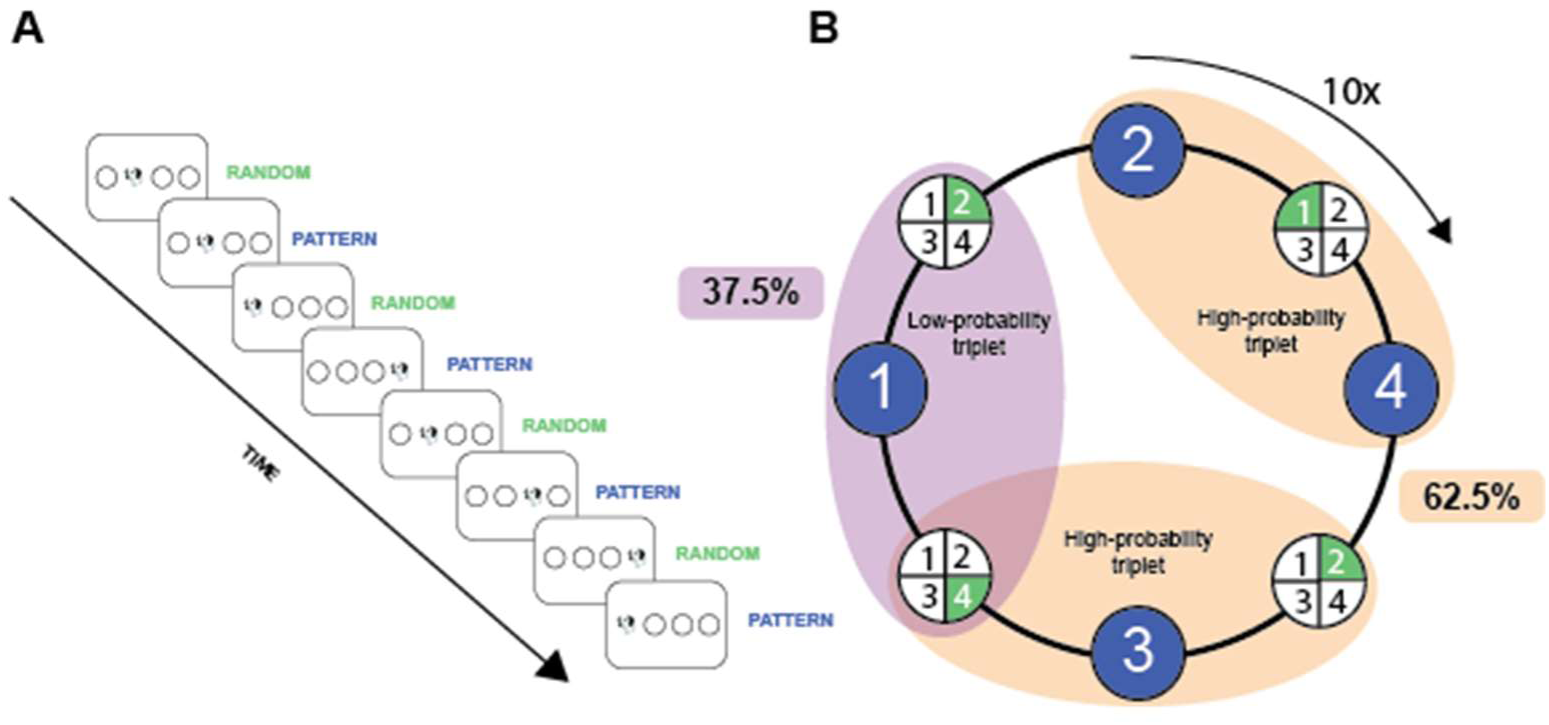
Structure of the ASRT. *Note.* A) The eight-element sequence consisted of alternating random and pattern stimuli. The first and every other element was randomly selected from four possible locations, and the remaining elements followed a fixed pattern (e.g., r 2 r 4 r 3 r 1, see Fig. 1A). The alternating sequence was repeated 10 times within a block. B) High-probability triplets occurred more frequently, as they could be formed both when a triplet started with a pattern-based stimulus and when it started with a random stimulus—together accounting for 62.5% of all triplet occurrences. In contrast, low-probability triplets could only occur when the triplet began with a random stimulus, resulting in a lower frequency (37.5%). Because high-probability triplets appear more frequently, participants are more likely to implicitly learn them over time. Therefore, the performance difference between high- and low-probability triplets serves as an index of habit learning.

The task starts with one practice block containing 80 random trials. Following the practice, participants completed 20 blocks of the ASRT task, with a short 15-minute break after the 15th block, during which they completed questionnaires. Each block consists of 10 repetitions of a randomly selected 8-element sequence, resulting in 1600 stimuli per participant in total. For analysis, the blocks were grouped into four epochs, each containing five blocks.

#### 2.2.7 Ten-Item Internet Gaming Disorder Test

Gaming Disorder risk (GDR) was assessed using a self-report measure, the Hungarian version of the Ten-Item Internet Gaming Test (IGDT-10; Király et al., 2017). This questionnaire is one of the most widely accepted and commonly used tools for assessing GDR, as it is based on the DSM-5 criteria and demonstrates good psychometric properties (Király et al., 2019; Yoon et al., 2021).

The IGDT-10 consists of ten items, each corresponding to one of the DSM-5 diagnostic criteria for IGD. The only exception is criterion 9, which refers to "jeopardizing or losing a significant relationship, job, or educational or career opportunity due to participation in internet games." This criterion was assessed using two separate items (items 9 and 10). However, to ensure consistency, responses to these two items were combined so that only a single point was assigned if the participant responded “often” to either or both items. Participants rated each item on a 3-point Likert scale (0 = never, 1 = sometimes, 2 = frequently). However, to align with the dichotomous scoring approach used in the DSM-5, responses of "never" and "sometimes" were coded as not meeting the criterion (0 points), while "frequently" was coded as meeting the criterion (1 point). Consequently, participants could achieve a maximum score of 9 points. Based on the DSM-5 classification system, individuals who met at least five criteria were categorized into the GDR group. It is important to emphasize that the IGDT-10 is not a diagnostic tool but rather a screening instrument intended to assess GD risk at a preliminary level. In the current sample, the questionnaire demonstrated adequate reliability with Cronbach’s alpha of 0.84.

### 2.3 Statistical analysis

Statistical analyses were conducted using JASP (Version 0.19.3.0; JASP Team, 2024) and IBM SPSS Statistics (Version 28; IBM Corp., 2021) for descriptive statistics and data cleaning. Group comparisons were performed in RStudio (Version 2024.12.0.467), while interaction analyses were conducted using RStudio (Version 2023.6.2.561). For data visualization, we used Python (Version 3.10.12) with the pandas, io, and matplotlib packages (Hunter, 2007; McKinney, 2010). We first examined whether the GDR, RG, and NG groups differed across sociodemographic variables. Participants were compared across gender, age, education, place of residence, current socioeconomic status (SES), and childhood SES. For continuous variables, independent sample t-tests or Mann-Whitney tests were conducted based on normality assumptions, while categorical variables were analyzed using Chi-squared tests. To account for potential confounding effects, sociodemographic variables that showed significant differences between groups were planned to be included as covariates in the analyses.

To meet the ANCOVA assumption that only covariates significantly associated with the dependent variable should be included, we first identified those sociodemographic variables that significantly differed across the GDR, RG, and NG groups in preliminary analyses (gender, age, and education). Each was tested in a separate linear regression to determine whether it significantly predicted the dependent variable. Covariates meeting this condition were included in the corresponding ANCOVA model. If none of the candidate covariates met this criterion, a one-way ANOVA was performed instead of ANCOVA.

In all models—ANCOVA or ANOVA—we systematically tested statistical assumptions. Residuals were tested for normality using the Shapiro–Wilk test. If the residuals violated the assumption of normality (p < .05), we applied a non-parametric method: either a rank-transformed ANCOVA (RANCOVA) or a Kruskal–Wallis test, depending on the model structure. When residuals were normally distributed, we tested for the homogeneity of variances using Levene’s test. If this assumption was violated, we used a robust permutation-based ANCOVA via the *lmp* function. If both assumptions were satisfied, a standard parametric ANCOVA (or ANOVA) was conducted. Partial eta squared was reported as the effect size for parametric models, and eta squared was computed for the Kruskal–Wallis tests.

When the overall group effect reached statistical significance in the parametric ANOVA or ANCOVA model, post hoc pairwise comparisons were performed using Bonferroni-corrected t-tests. In cases where assumptions of parametric testing were violated and a non-parametric method was used instead, Dunn’s post hoc tests with Bonferroni correction were applied. This approach ensured valid and conservative group-level inferences across all analytical pathways.

For the evaluation of the ASRT task, we computed scores by calculating the median reaction time (RT) and mean accuracy for each epoch (consisting of 5 blocks, each with 80 stimuli) and each participant (see Supplementary Materials S2 for details). To analyze the habitual learning process, we conducted two mixed-design analyses of variance (ANOVAs), with accuracy and RT as the dependent variables. First, we checked whether the sociodemographic variables that differed between groups influenced performance by conducting linear regression analyses with accuracy and RT as outcomes. Variables that significantly predicted performance were included as covariates in the ANOVA models. The three-level grouping variable included the GDR, RG, and NG groups. The within-subject factors were triplet type (two levels: high-probability and low-probability) and epoch (four levels). Sphericity was assessed using Mauchly’s test, and the Greenhouse–Geisser correction was applied when the assumption of sphericity was violated.

To identify the latent structure of our executive function (EF) measures, we conducted exploratory factor analyses (EFA) using maximum likelihood estimation with varimax rotation. Prior to analysis, we ensured data suitability through standard diagnostics (e.g., KMO, Bartlett’s test; details in Supplementary Materials S4). The number of factors to retain was determined via Horn’s parallel analysis using a conservative criterion. Model fit was assessed using RMSEA and SRMR, and individual factor scores were computed using regression-based methods. These factor scores were then compared across groups, following the same statistical assumptions and checks as described above.

To examine the interaction between goal-directed and habitual processes, we conducted generalized linear mixed model (GLMM) analyses, performed in R 4.2.1. A detailed description of the analytical approach, as well as the full results, are provided in Supplementary Materials S5. Models were fit in R using the *afex* package (Singmann et al., 2024), with reaction times (RTs) analyzed using linear mixed models on log-transformed data, and accuracy using logistic mixed models. Triplet type, Epoch, Group, and EF1 and EF2 factor scores were included as fixed effects, along with relevant covariates (Age, Education, Gender). Following current best practices, models were initially specified with a maximal random effects structure and simplified only in cases of convergence issues.

Model assumptions (e.g., linearity, homoscedasticity) were assessed and largely met. Fixed effects were tested using Type III tests (RTs) or likelihood ratio tests (accuracy), and significant interactions with continuous predictors were probed at ±1 SD. Marginal and conditional R² values, ICCs, and random effect estimates are reported, alongside corrected and uncorrected p-values for post hoc comparisons (Šidák-adjusted where applicable). Effect sizes are interpreted according to standard conventions for individual differences research. α = .05 was used for all inferential tests. All data and code necessary to reproduce the results of this study are available on the OSF platform (see Data Availability Statement).

## 3. Results

### 3.1 Sociodemographic variables

When comparing the groups, we found significant differences in age, gender, and level of education. Regarding age, both the RG and GDR groups were significantly younger than the NG group, and the GDR group was also younger than the RG group. In terms of gender, the proportion of female participants was highest in the NG group (58.54%) and lower in the RG (21.42%) and GDR (25.81%) groups. Regarding the level of education, a larger proportion of participants held a college or university degree in the NG group (82.93%) compared to the RG (59.52%) and GDR (40%) groups, while the proportion of participants with only a high school diploma was highest in the GDR group (50%), followed by the RG group (30.95%) and the NG group (9.76%). However, no significant group differences were observed in terms of place of residence or current and childhood socioeconomic status (SES) (see Supplementary Materials S3 for details).

Consequently, age, gender, and education were considered as control variables in the analyses. To assess whether they should be included as covariates in the subsequent ANCOVA models, we examined their associations with the dependent variables through linear regression analyses. Details on which of the three control variables were included for each score can be found in the Supplementary Materials S3.

### 3.2 Working memory

To assess working memory capacity, we compared the performance of the GDR, RG, and NG groups on the DSPAN, CSPAN, and n-back tasks, while controlling age, gender, and level of education. Significant group differences were observed on the DSPAN task, with the GDR group performing significantly worse than the NG group. No significant differences were found between the RG and NG groups or between the GDR and RG groups on the DSPAN task. Similarly, significant group differences were observed on the CSPAN task, which measured complex working memory capacity. The GDR group again performed significantly worse than the NG group and also showed significantly lower performance compared to the RG group. In contrast, no significant differences were observed between the RG and NG groups. For further details, see Table 2 and Fig. 2.

**Fig. 2.**
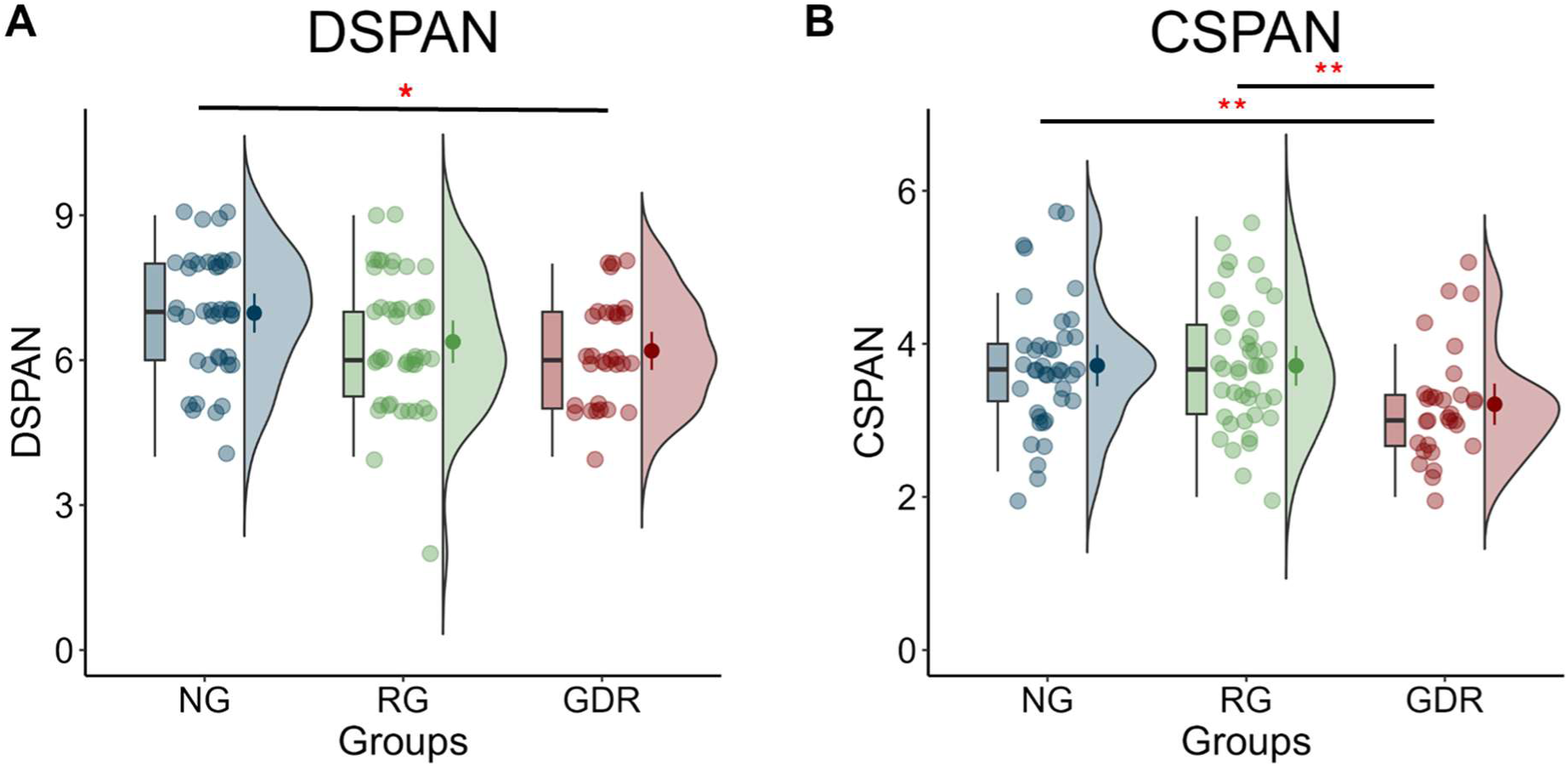
Digit Span and Counting Span (CSPAN; B) scores of the non-gamer (NG), recreational gamer (RG), and gaming disorder risk (GDR) groups. *Note.* This figure includes (A) the Digit Span (DSPAN) and (B) Counting Span (CSPAN) scores (* *p* < .05; ** *p* < .01)

**Table 2.**
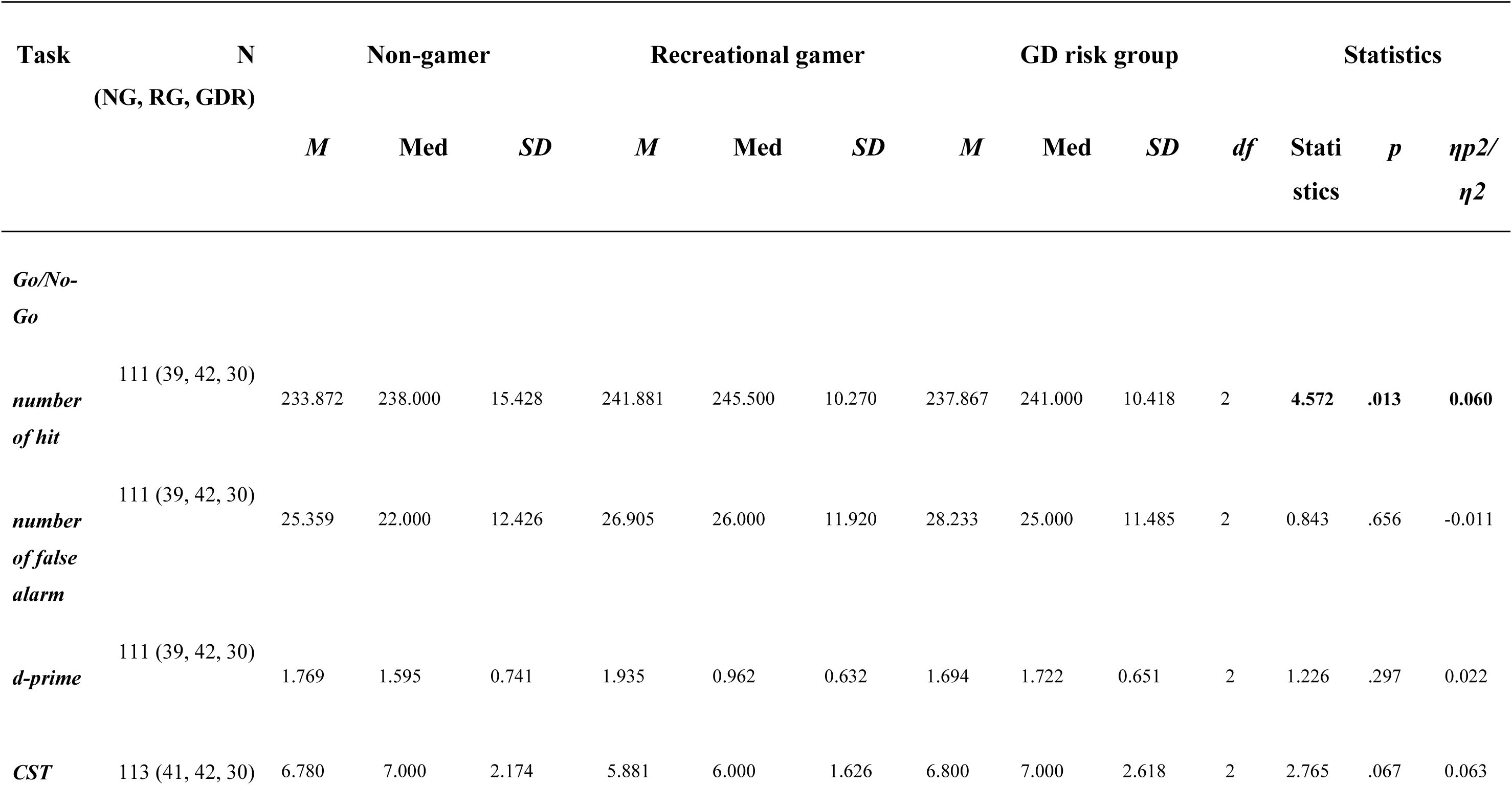
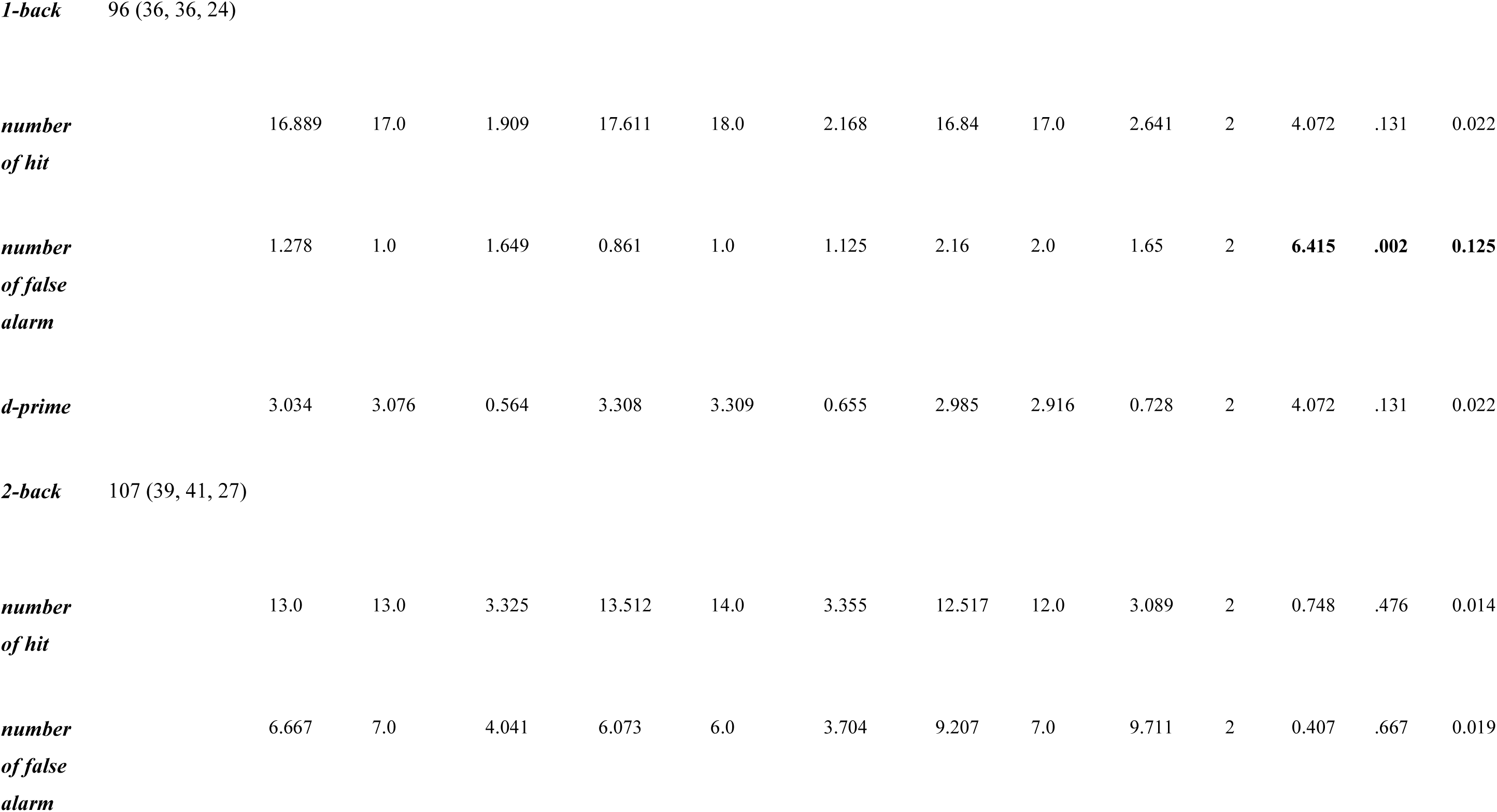
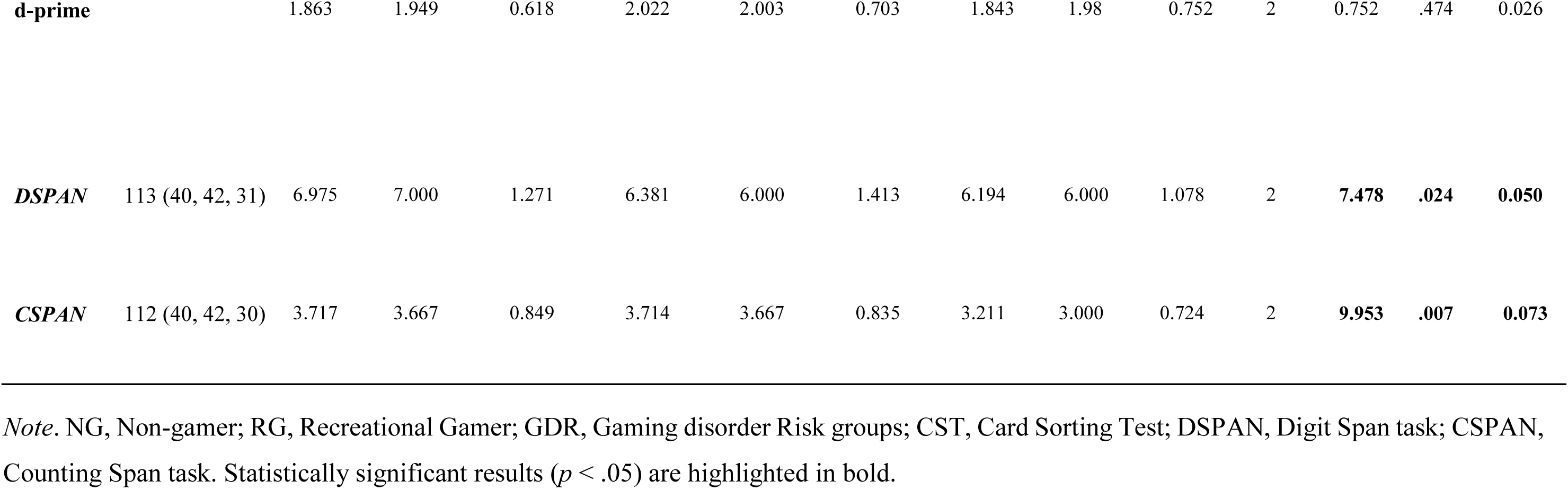
ANCOVA results of executive function scores across groups adjusted for age, gender, and education.

In the 1-back task, there was a significant difference in the number of false alarms, with the GDR group exhibiting more false alarms compared to the RG group. On a trend level, the GDR group also showed more false alarms compared to the NG group. However, there was no significant difference in false alarms between the RG and NG groups. Additionally, there was a trend level difference in the d-prime score between the three groups. The analysis revealed no significant group differences in d-prime scores, number of hits, false alarms, or reaction times in the 2-back task (see Table 1, Fig. 3).

**Fig. 3.**
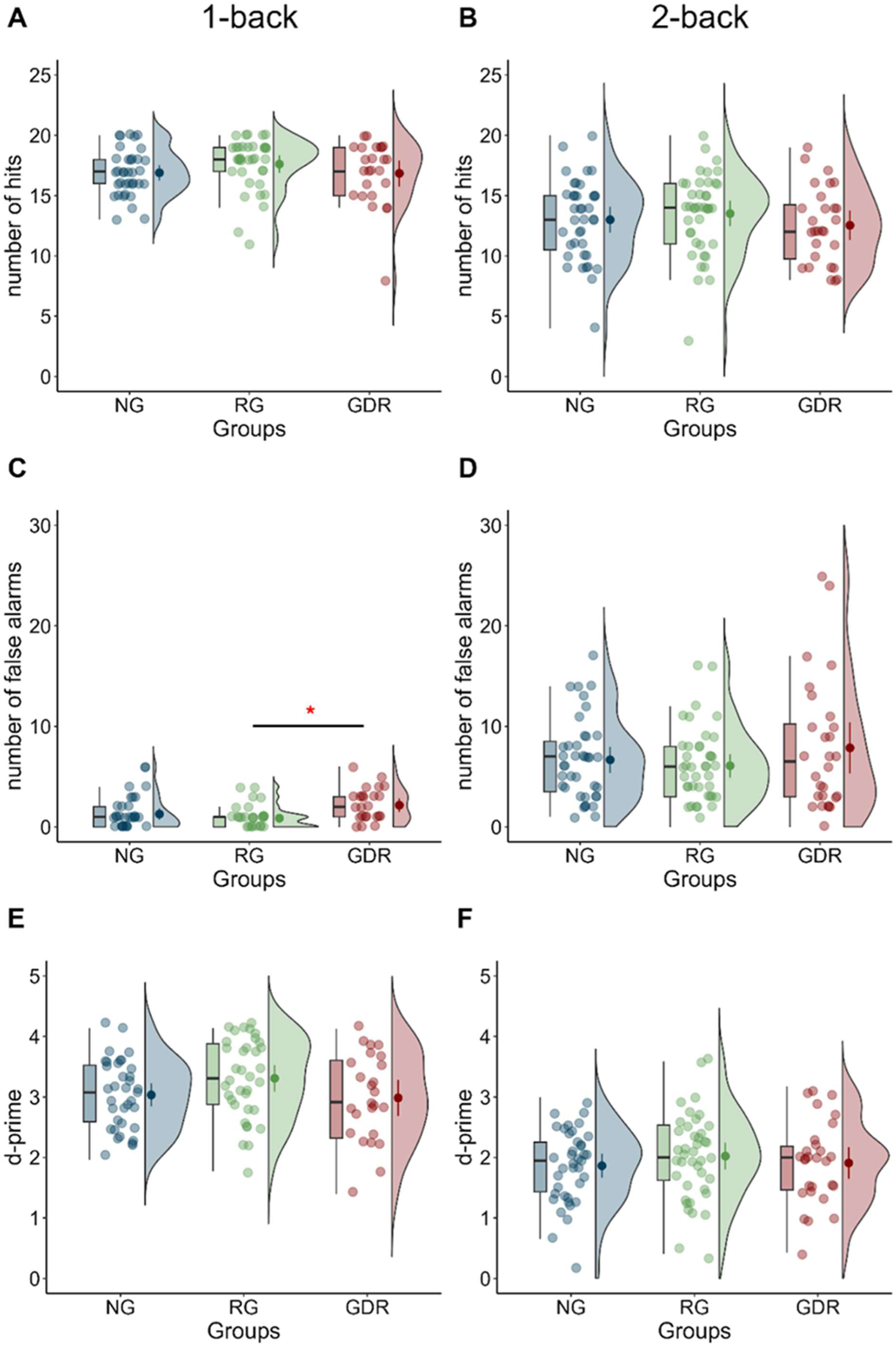
N-back scores of the non-gamer (NG), recreational gamer (RG), and gaming disorder risk (GDR) groups. *Note*. In the first column, the figures display the results of the 1-back test: (A) the number of hits, (C) the number of false alarms, and (E) the d-prime scores. The second column shows the scores for the 2-back test: (B) the number of hits, (D) the number of false alarms, and (F) the d-prime scores (\*\**p* < .01).

### 3.3 Inhibitory control

The Go/No-Go task, designed to assess inhibitory control, revealed significant group differences in the number of hits across the three groups. The RG group outperformed the NG group, responding correctly to more target stimuli. No significant differences were observed between the NG and GDR groups or between the GDR and RG groups. Additionally, no significant differences were found among the three groups in the discriminability or false alarm scores (see Table 2, Fig. 4A).

**Fig. 4.**
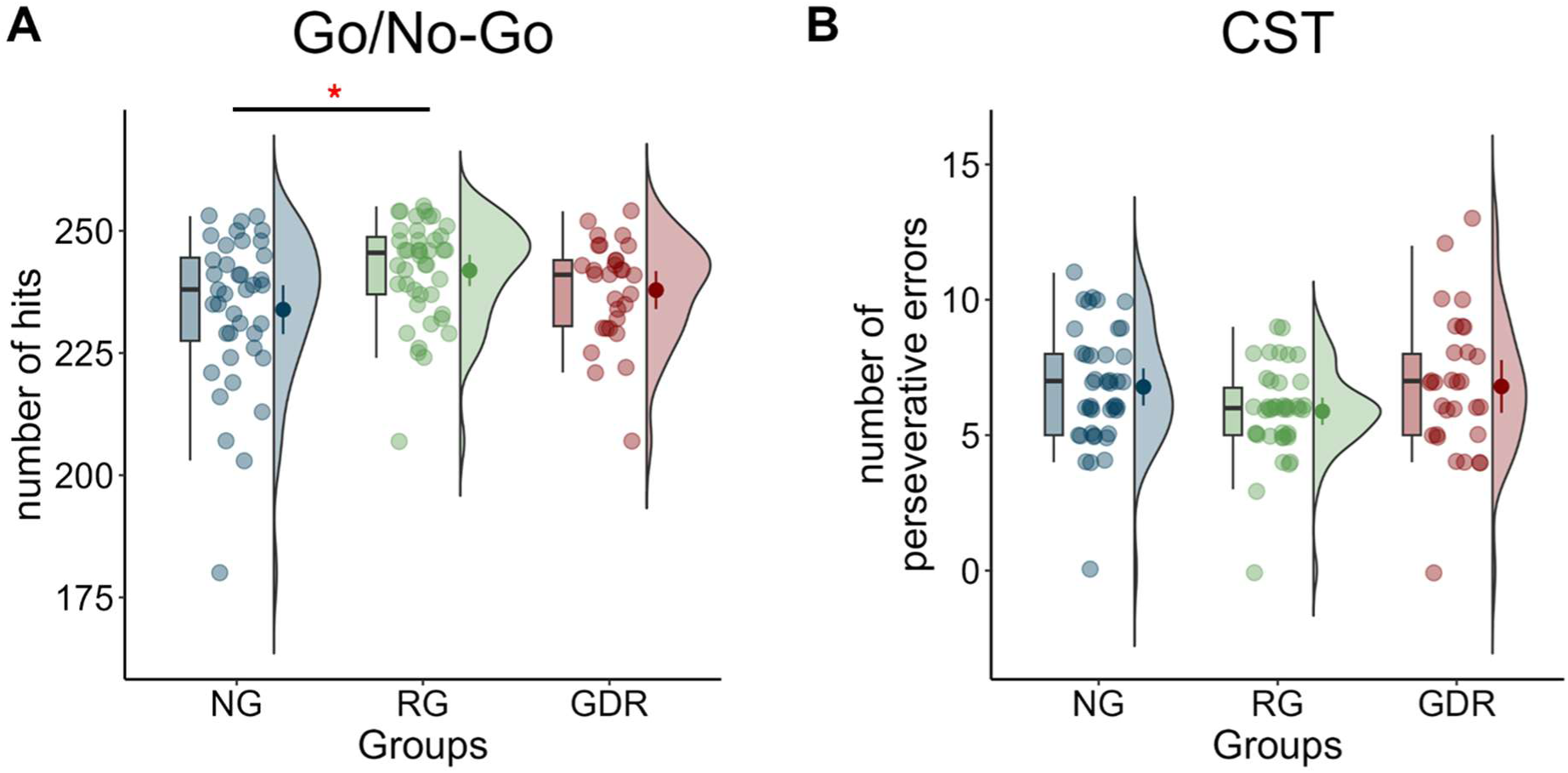
Cognitive flexibility and inhibitory control scores of the non-gamer (NG), recreational gamer (RG), and gaming disorder risk (GDR) groups. *Note.* This figure includes (A) the number of hits in the Go/No-Go task and (B) the number of perseverative errors in the Card Sorting Task (CST) (* *p* < .05).

### 3.4 Cognitive flexibility

To assess cognitive flexibility, we used the CST task, which revealed trend-level differences among the three groups, although it did not reach significance (see Table 2, Fig. 4B). The RG group exhibited the fewest perseverative errors, followed by the NG group, while the GDR group showed the highest number of perseverative errors.

### 3.5 Habit learning

We performed a mixed-design ANOVA to examine group differences in habit learning based on reaction times and accuracy. Please note that the main effects and interactions involving the Triplet Type factor indicate differences in habit learning, while those excluding this factor reflect overall reaction time differences.

Age, gender, and level of education significantly predicted both reaction time (RT) and accuracy in preliminary linear regression analyses; therefore, all three variables were included as covariates in the final models. The mixed-design ANOVA on RT revealed a marginal main effect of Epoch, indicating a trend toward improved reaction times across the task. In addition, a significant main effect of Group was observed, suggesting overall differences in RT between the groups. Post hoc pairwise comparisons (Bonferroni-adjusted) showed that the recreational gaming (RG) group responded significantly faster (*M* = 364 ms) than the non-gaming (NG) group (*M* = 391 ms), *p* = .027. No significant differences emerged between the other group pairs. Moreover, the analysis revealed a significant Group × Epoch interaction, indicating that the rate of performance improvement varied across groups over time. Among the covariates, age showed a robust main effect, with older participants generally exhibiting slower reaction times. A significant Age × Epoch interaction, further indicated that age influenced how performance changed over time. In contrast, the Education × Epoch interaction was not significant, and no other covariate-related interactions reached significance. Finally, although the main effect of Triplet Type was significant, —indicating faster responses to high-probability triplets—there was no significant Group × Triplet Type interaction. This suggests that, after controlling for covariates, the three groups did not differ in their sensitivity to the underlying triplet structure, reflecting comparable levels of habitual learning (for further details, see Table 3, Fig. 5).

**Fig. 5.**
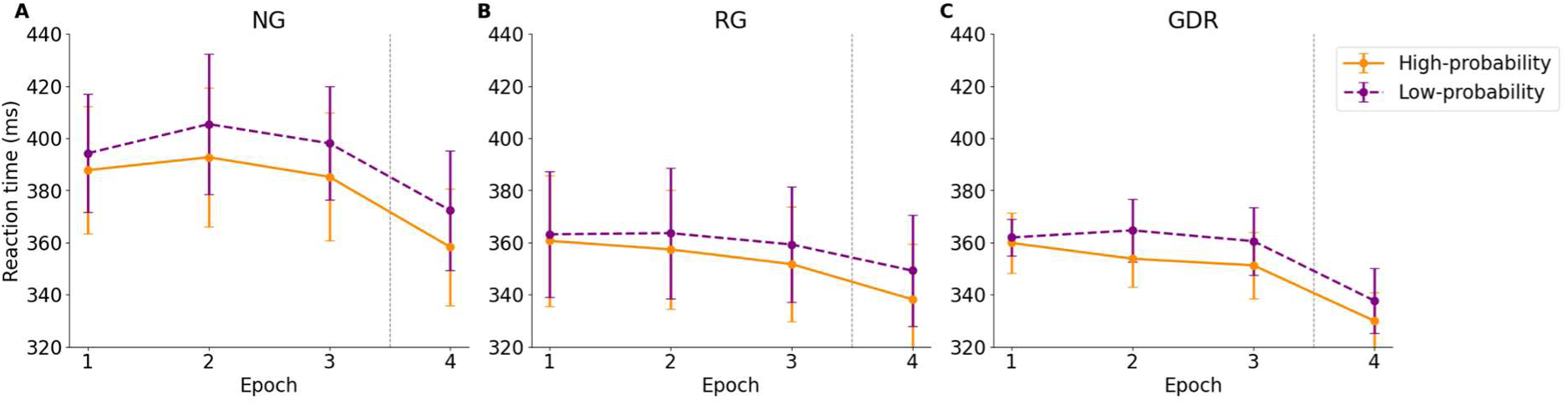
Reaction time in the Alternating Serial Reaction Time (ASRT) task of the non-gamer (NG), recreational gamer (RG), and gaming disorder risk (GDR) groups. *Note*. The figure illustrates reaction time (RT) data on the ASRT task. The orange and purple lines correspond to RTs for high-probability and low-probability triplets, respectively. A greater separation between these lines indicates stronger statistical learning. The gray dashed line marks a 20-minute break in the procedure. Error bars represent 95% confidence intervals.

**Table 3.** Results of the mixed-design ANOVA on the reaction time of the Alternating Serial Reaction Time task.

| Effect | *df* | *F* | *p* | $\eta^2p$ |
| --- | --- | --- | --- | --- |
| **Group** | **2, 86** | **3.98** | **.022** | **.085** |
| **Age** | **1, 86** | **31.49** | **<.001** | **.268** |
| Gender | 1, 86 | 2.17 | .145 | .025 |
| Education | 4, 86 | 1.64 | .172 | .071 |
| **Triplet Type** | **1, 86** | **10.84** | **.001** | **.112** |
| Group × Triplet Type | 2, 86 | 1.50 | .229 | .034 |
| Age × Triplet Type | 1, 86 | 0.25 | .617 | .003 |
| Gender × Triplet Type | 1, 86 | 2.95 | .090 | .033 |
| Education × Triplet Type | 4, 86 | 0.99 | .419 | .044 |
| Epoch | 2.54, 218.13 | 2.24 | .095 | .025 |
| **Group × Epoch** | **5.07, 218.13** | **3.60** | **.004** | **.077** |
| **Age × Epoch** | **2.54, 218.13** | **2.87** | **.046** | **.032** |
| Gender × Epoch | 2.54, 218.13 | 2.47 | .073 | .028 |
| Education × Epoch | 10.15, 218.13 | 1.40 | .180 | .061 |
| Triplet Type × Epoch | 2.97, 255.49 | 1.00 | .393 | .011 |
| Group × Triplet Type × Epoch | 5.94, 255.49 | 1.06 | .386 | .024 |
| Age × Triplet Type × Epoch | 2.97, 255.49 | 1.32 | .267 | .015 |
| Gender × Triplet Type × Epoch | 2.97, 255.49 | 0.21 | .885 | .002 |
| Education × Triplet Type × Epoch | 11.88, 255.49 | 0.89 | .554 | .040 |

*Note.* Values with statistical significance at $p < .05$ are highlighted in bold.

In the analysis of accuracy, age, gender, and education were included as covariates, as they significantly predicted accuracy in preliminary regression analyses. The ANOVA revealed a significant main effect of Epoch, indicating that participants’ accuracy changed over time. In addition, there was a significant main effect of Triplet Type, demonstrating that participants, overall, showed evidence of habit learning, with higher accuracy on high-probability triplets compared to low-probability ones. Among the covariates, age emerged as a significant predictor, suggesting that performance was influenced by participants’ age. No other main effects or interactions have reached significance, including those involving the Group variable. Therefore, we conclude that the three groups did not differ significantly in their habit learning performance after controlling for covariates (see Table 4, Fig. 6).

**Fig. 6.**
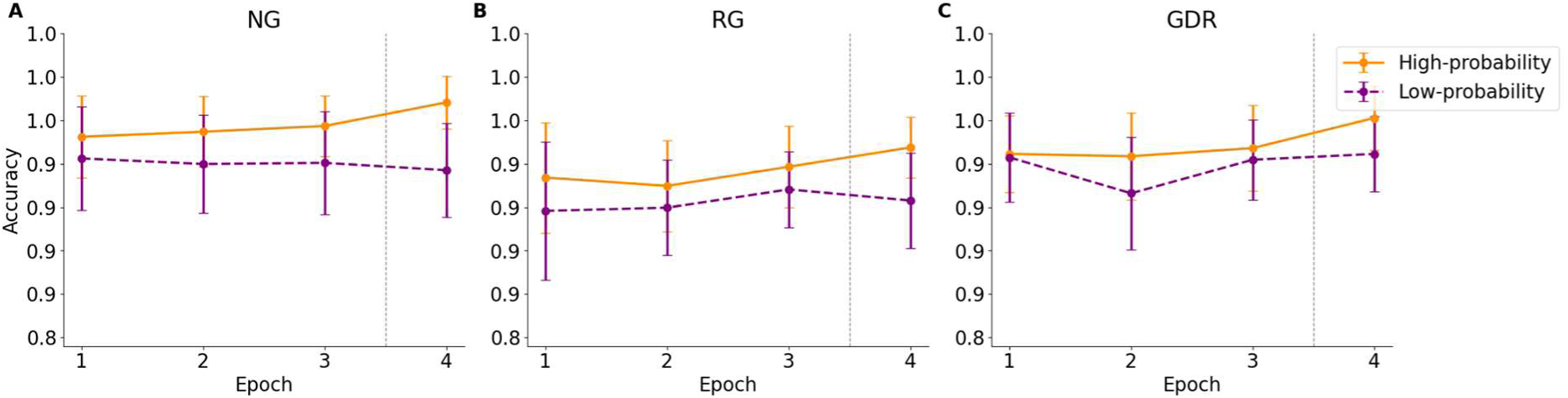
Accuracy in the Alternating Serial Reaction Time (ASRT) task of the non-gamer (NG), recreational gamer (RG), and gaming disorder risk (GDR) groups. *Note*. The figure illustrates accuracy (ACC) on the ASRT task. The orange and purple lines correspond to RTs for high-probability and low-probability triplets, respectively. A greater separation between these lines indicates stronger statistical learning. The gray dashed line marks a 20-minute break in the procedure. Error bars represent 95% confidence intervals.

**Table 4.** Results of the mixed-design ANOVA on accuracy of the Alternating Serial Reaction Time task.

| Effect | *F* | *df* | *p* | $\eta^2p$ |
| --- | --- | --- | --- | --- |
| Group | 1.00 | 2, 87 | .370 | .023 |
| **Age** | **9.10** | **1, 87** | **.003** | **.095** |
| Gender | 1.21 | 1, 87 | .274 | .014 |
| Education | 0.95 | 4, 87 | .438 | .042 |
| **Triplet Type** | **11.39** | **1, 87** | **.001** | **.116** |
| Group × Triplet Type | 0.73 | 2, 87 | .485 | .017 |
| Age × Triplet Type | 1.90 | 1, 87 | .172 | .021 |
| Gender × Triplet Type | 0.55 | 1, 87 | .461 | .006 |
| Education × Triplet Type | 0.45 | 4, 87 | .772 | .020 |
| **Epoch** | **4.91** | **2.39, 207.91** | **.005** | **.053** |
| Group × Epoch | 0.66 | 4.78, 207.91 | .649 | .015 |
| Age × Epoch | 1.54 | 2.39, 207.91 | .213 | .017 |
| Gender × Epoch | 1.29 | 2.39, 207.91 | .280 | .015 |
| Education × Epoch | 1.39 | 9.56, 207.91 | .188 | .060 |
| Triplet Type × Epoch | 1.37 | 2.83, 246.06 | .254 | .016 |
| Group × Triplet Type × Epoch | 1.06 | 5.66, 246.06 | .388 | .024 |
| Age × Triplet Type × Epoch | 1.56 | 2.83, 246.06 | .202 | .018 |
| Gender × Triplet Type × Epoch | 0.19 | 2.83, 246.06 | .892 | .002 |
| Education × Triplet Type × Epoch | 1.02 | 11.31, 246.06 | .431 | .045 |

*Note.* Values with statistical significance at $p < .05$ are highlighted in bold.

Fig. 7 illustrates the distinct cognitive profiles of the NG, RG, and GDR groups across multiple tasks.

**Fig. 7.**
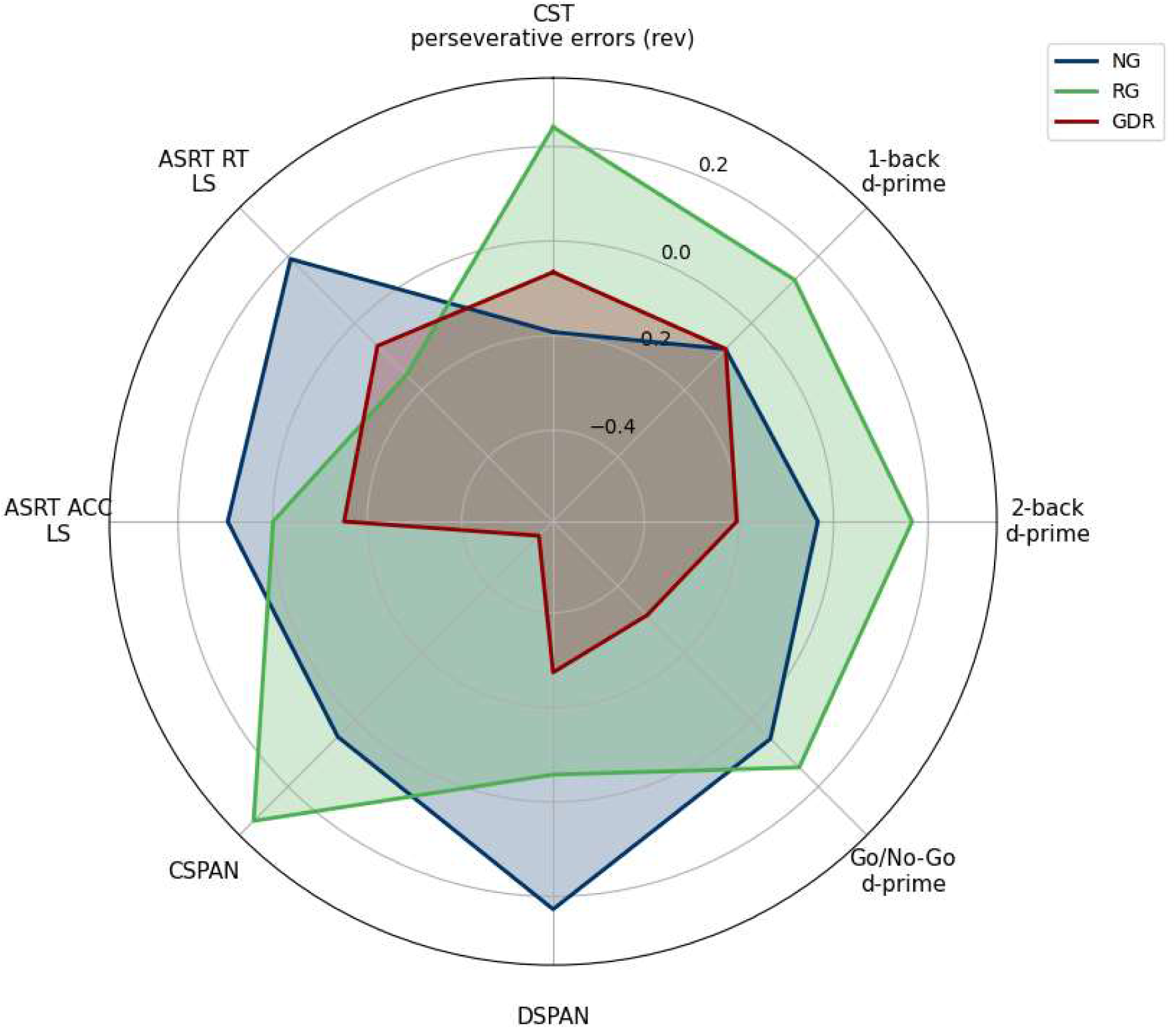
Cognitive profiles of the non-gamer (NG), recreational gamer (RG), and gaming disorder risk (GDR) groups. *Note.* This radar plot displays standardized residual scores from various cognitive tasks, including the Card Sorting Test (CST), 1-back (d-prime), 2-back (d-prime), Go/No-Go (d-prime), Digit Span (DSPAN), Complex Span (CSPAN), and learning scores from the Alternating Serial Reaction Time task (ASRT): Accuracy Learning Score (ACC LS) and Reaction Time Learning Score (RT LS). The plot compares cognitive profiles of non-gamers (NG), recreational gamers (RG), and individuals at risk for Gaming Disorder (GDR).

For the CST, the number of perseverative errors was used as the performance indicator. As lower error counts indicate better performance, these values were reverse-coded so that higher values consistently reflect better cognitive functioning across all axes.

To control for potential confounding effects, each variable was regressed on age, gender, and education level using ordinary least squares (OLS) regression. The resulting residuals—i.e., the portion of task performance unexplained by demographic factors—were then averaged within each group and used in the radar plot. This approach ensures that the visualized group differences reflect task-specific cognitive performance, independent of demographic characteristics.

### 3.6 Factor analysis of executive function tasks

A detailed factor analysis of executive function (EF) tasks is presented in Supplementary Materials S4. Briefly, parallel analysis supported a two-factor solution, explaining 35% of the total variance. The first factor reflected simple working memory capacity (CSPAN, DSPAN), while the second factor was primarily associated with complex working memory and inhibition tasks (1-back, 2-back, Go/No-Go). CST did not load strongly on either factor. Factor scores derived from this solution were used in the subsequent analyses (see details in Table 5 and Supplementary Materials S4).

**Table 5.** Factor loadings and communalities of each EF measure in the two factor varimax rotated solution.

|  | Factor 1 | Factor 2 | Communality |
| --- | --- | --- | --- |
| CST | 0.27 | 0.02 | 0.06 |
| 1-back | 0.04 | **0.46** | 0.20 |
| 2-back | 0.28 | **0.52** | 0.37 |
| Go / No-Go | 0.01 | **0.70** | 0.47 |
| CSPAN | **0.98** | 0.19 | 0.87 |
| DSPAN | **0.32** | 0.04 | 0.14 |

*Note.* Loadings above 0.30 are highlighted in bold to aid interpretation.

### 3.7 Comparison of executive function factors between the groups

For EF1, which was primarily loaded on CSPAN, DSPAN, and 2-back tasks, a significant group difference was observed. Specifically, the GDR group performed significantly worse on this factor compared to both the NG and RG groups, while no significant difference was found between the NG and RG groups. In contrast, no significant group differences emerged for EF2, which was primarily associated with the Go/No-Go, 1-back, and 2-back tasks (see details in Table 2).

### 3.8 Habit learning and executive functions in the three groups

The detailed results of the GLMMs examining the interaction between habit learning and executive functions across the three groups are presented in Supplementary Materials S5. In summary, in the models using RT-based habit learning scores, the results suggest that overall visuomotor performance was comparable between the groups, and that competition between EF2 and habit learning was stronger in the RG group than in either the NG or GDR groups. In the models using accuracy-based habit learning scores, EF1 scores were unrelated to overall accuracy. Instead, they were positively associated with habit learning ability in the NG and GDR groups, with no such association in the RG group. EF2 scores were associated with better overall accuracy only in the GDR group and showed a generally negative relationship with habit learning across all groups.

## 4. Discussion

Although many individuals can regulate their gaming habits, some experience such a profound loss of control that it becomes a central issue in their lives, manifesting as behavioral addiction. To better understand the cognitive mechanisms underlying these differing gaming habits, we examined executive functions, habit learning, and how these processes interact. Our findings revealed selective executive function impairments in individuals at risk for gaming disorder, particularly reduced working memory capacity and more impulsive response tendencies, reflected in higher false alarm rates. In contrast, recreational gamers showed enhanced attention control and faster responses without sacrificing accuracy, suggesting potential cognitive benefits of non-problematic gaming. No significant group differences emerged in working memory updating, inhibitory control, or cognitive flexibility, and habit learning appeared comparable across groups. However, the relationship between executive control and habit learning varied across groups. To the best of our knowledge, this is the first study to compare comprehensive cognitive profiles—including both executive functioning and habit learning, as well as their interaction—across non-gamers, recreational gamers, and individuals at risk for gaming disorder.

### 4.1 Executive functions

In line with our hypotheses, the gaming disorder risk group performed significantly worse than the non-gamer group on the short-term memory task, and worse on the complex working memory task than the other two groups. Importantly, these tasks primarily assess the storage function of working memory (Case et al., 1982; Racsmány et al., 2006). In tasks measuring the updating component, such as the 1-back and the 2-back tasks, no group differences were found (Ragland et al., 2002). These findings indicate that gaming disorder is associated with decreased working memory capacity, specifically related to storage processes. This result aligns with previous research summarized in the review by Ngetich et al. (2023), emphasizing working memory impairments in gaming disorder. The lack of differences between recreational gamers and non-gamers suggests that gaming, in the absence of addiction, possibly is not related to impaired working memory performance. These findings are particularly intriguing given that impairments in working memory have been observed in gambling disorder, compulsive buying disorder and work addiction as well (Berta et al., 2023; Derbyshire et al., 2014; Ngetich et al., 2023). This suggests that this domain-specific cognitive deficit may play an important role in behavior addictions. Since working memory is a key component of higher-order executive functions—such as planning and problem-solving— deficits in this domain may contribute to the maintenance of maladaptive behavior patterns and difficulties in self-regulation (Collins & Koechlin, 2012; Diamond, 2013).

Although no significant group differences were found in overall performance on the n-back tasks, the analysis of error types revealed a more nuanced picture. Specifically, performance on the 1-back task showed a higher number of false alarms in the gaming disorder risk group compared to the recreational gamer group. Although this is a relatively simple working memory task, a high false alarm rate without a correspondingly high hit rate may reflect increased behavioral impulsivity (Bezdjian et al., 2009). This finding is especially relevant in light of the potential dual nature of gaming effects: while gaming may enhance stimulus detection and response accuracy, gaming disorder may be accompanied by more impulsive response styles that counteract these benefits. This pattern may also be important considering that such impulsive behavior and lack of self-control could contribute to the difficulty in resisting the urge to play (Warburton et al., 2022).

Contrary to our initial hypothesis, no significant differences were observed between the groups in inhibitory control, as assessed by the d-prime score in the Go/No-Go task (Hinton, 2015). This finding contradicts previous research, which has consistently shown a link between gaming disorder and impaired inhibitory control (Argyriou et al., 2017). Notably, impairments in inhibitory control may be more pronounced with addiction-specific stimuli due to attentional bias towards gaming-related cues, which may explain the lack of significant findings with neutral stimuli in the present task (Lorenz et al., 2013; Maleki et al., 2024; Yao et al., 2015). These considerations underscore the importance of incorporating both neutral and addiction-specific stimuli (e.g., images from video games) within the same paradigm in future studies to more accurately capture context-dependent impairments in inhibitory control. It is also important to note that, since most cognitive tasks are performed on computers, the testing environment itself may serve as a game-related cue—particularly for individuals at risk for gaming disorder. Therefore, identifying alternative methods, such as virtual reality (VR), could help mitigate attentional biases triggered by addiction-related cues and also provide a more ecologically valid environment for assessing executive functions.

Although no significant differences were observed in the Go/No-Go’s d-prime score, a notable difference was found in the hit rate, with the recreational gaming group demonstrating a significantly higher rate compared to the non-gamer group. This result, while not conclusive for superior inhibitory functioning, may suggest enhanced response readiness and attentional control in the recreational gaming group (Eimer, 1993). This finding aligns with prior research suggesting that video game play can improve attentional processing and visuospatial abilities (Stanmore et al., 2017). An alternative explanation is that video games may not enhance cognitive abilities per se, but rather individuals with higher baseline cognitive skills are more likely to engage in gaming (Waris et al., 2019). Their superior performance may lead to more frequent positive reinforcement during gaming, which in turn increases their motivation to continue playing. However, it is interesting that this result did not appear in the gaming disorder risk group, who also have gaming experience, suggesting that gaming experience alone may not account for the differences observed. Instead, these findings point to the possibility that the cognitive benefits of video gaming—such as enhanced attentional control—may be present only in the context of non-problematic use and may diminish or disappear when gaming becomes problematic. It is also possible that in cases where gaming becomes problematic, other factors—such as sleep deprivation, or stress resulting from conflicts related to gaming—may negatively impact inhibitory functions (Choong et al., 2025; Roos et al., 2017). While these explanations remain speculative, clarifying the direction and causality of these relationships would require longitudinal studies to better understand how cognitive functioning evolves in relation to gaming behavior over time.

Contrary to our initial hypothesis, the results of the Card Sorting Test showed no significant differences in cognitive flexibility between the groups, suggesting that this executive function may not be linked to either recreational gaming or gaming disorder. While the recreational gaming group exhibited the fewest perseverative errors, this trend did not reach statistical significance, indicating a subtle pattern that may merit further investigation. These findings are particularly interesting in light of previous studies that reported reduced cognitive flexibility in individuals with gaming disorder (Jeong et al., 2016; Jiang et al., 2020) relied on clinically diagnosed samples. In contrast, one study using self-reported questionnaires (Ryu et al., 2021) found no behavioral-level differences in cognitive flexibility—only neural-level alterations. While it is possible that cognitive flexibility is a relatively preserved function in individuals with gaming disorder, these inconsistencies also point to the potential moderating role of clinical severity and diagnostic approach. Therefore, future studies should further explore whether impairments in cognitive flexibility are more pronounced in clinically diagnosed populations or under more demanding task conditions.

Following the analysis of group differences at the task level, we also performed a factor analysis to gain a deeper understanding of the broader role of executive functions. This analysis revealed two overarching factors. The first factor encompassed the DSPAN and CSPAN tasks, while the second factor included the 1-back, 2-back, and Go/No-Go tasks. The Card Sorting test did not load onto either of these factors. Consistent with the task-level findings, we observed group differences in the first factor, which is more closely related to storage functions. In this factor, the high-risk gaming disorder group performed the worst, significantly differing from both the non-gamer and recreational-gaming groups, who performed similarly. On the other hand, the second factor, which is more closely associated with updating and inhibition, did not show any significant group differences. These results are in line with the task-specific findings, suggesting that gaming disorder is most strongly associated with impaired working memory capacity, particularly regarding storage functions.

### 4.2 Habit learning

Contrary to our hypotheses, neither gaming disorder nor recreational gaming was found to be associated with enhanced habit learning. Though Romano Bergstrom et al. (2012) found improved sequence learning on the ASRT task among video game players, their sample was limited to university students, which may indicate that gaming’s benefits on habit learning are more pronounced in younger populations. Notably, their educational context and associated life circumstances might have facilitated the emergence of the effects they observed. In our sample, despite the absence of enhanced habit learning in the recreational gaming group, they exhibited significantly faster reaction times independently from sequence learning without the decline of accuracy, suggesting potential benefits in attentional and visuomotor processing (Stanmore et al., 2017). This finding is consistent with our results from the Go/No-Go task, where recreational gaming was linked to improved attentional control, supporting the idea that recreational gamers may experience advantages in attentional abilities and processing speed.

Interestingly, we did not observe enhanced habit learning in the gaming disorder risk group, despite previous research suggesting an over-reliance on habitual processes in individuals with gaming disorder (Zhou et al., 2021). Notably, these earlier findings were based on instrumental learning tasks, which capture different aspects of habit formation. In contrast, the ASRT task used in the present study measures implicit sequence learning—a specific facet of habitual functioning—which, according to our results, may remain intact in individuals with gaming disorder (Horváth et al., 2022). This is consistent with findings from work addiction, where no enhancement in habit learning was observed despite the presumed role of habit-oriented processes (Pesthy et al., 2025) and may suggest that this pattern extends to other behavioral addictions as well. Our results challenge models that propose a simple, monolithic "habitual system" that is uniformly enhanced in addiction.

### 4.3 Interaction between executive function factors and habit learning

Previous studies have already examined the imbalance between goal-directed and habitual processes in the context of gaming disorder (Kwon et al., 2024; Lei et al., 2025); however, our investigation of their competition between these processes—along with the inclusion of recreational gaming group—represents a novel approach that has not been explored before. In order to examine the relationship between these processes, we investigated how the two executive function-related factors—EF1 and EF2—are linked to habit learning scores across the three groups. No significant group differences were observed in overall habit learning performance, but the analysis of the two cognitive systems revealed several noteworthy findings, highlighting the complex and group-specific interactions between goal-directed and habitual processes.

In line with competition theory, we found a negative relationship between habit learning and EF2—the factor primarily loading on inhibitory control and the updating component of working memory—across all three groups, regardless of gaming habits or risk for behavioral addiction. This competitive interaction suggests that when prefrontal cortex–related goal-directed processes are attenuated, habit-based mechanisms become more dominant. A similar pattern was reported by Virag et al. (2015), who also found a negative association between goal-directed and habitual systems in both healthy controls and individuals with alcohol dependence, further supporting the idea that this antagonistic relationship is a generalizable feature of cognitive functioning. It is worth noting that these competitive patterns appeared only in models that used the habit learning score based on reaction time. In the case of accuracy, a negative relationship with habit learning was only found among recreational gamers, and only at certain points during the task. This suggests that accuracy- and reaction time-based measures in the ASRT task may reflect different cognitive processes (Janacsek et al., 2012; Kiss et al., 2022; Mulder & Van Maanen, 2013).

Interestingly, for the other factor which comprises tasks measuring working memory storage, a positive association between habit learning and EF1 emerged specifically in the non-gaming and gaming disorder risk groups, but not in the recreational gaming group. This raises the question of why both non-gamers and individuals at risk of gaming disorder showed a similar positive association between working memory storage and habit learning, even though group comparisons revealed significantly lower EF1 performance among those at gaming disorder risk. This discrepancy suggests that, although the relationship between cognitive systems appears similar, their underlying functioning may differ—possibly due to the engagement of different compensatory mechanisms.

One potential explanation is that working memory capacity, particularly its role in attentional resource allocation and the detection of recurring patterns, may facilitate habit learning under certain conditions. Although working memory is typically thought to aid performance in tasks with explicit learning components (Janacsek & Nemeth, 2013), previous research has shown that it may also contribute to learning in predominantly implicit contexts (Bo et al., 2012)—a possibility consistent with our findings. From this perspective, individuals with higher working memory capacity in both the non-gamer and gaming disorder risk groups may have been better able to allocate cognitive resources, thereby supporting improved task performance. These findings align with the suggestion by (Pedraza et al., 2024) that it is not working memory capacity per se, but rather the updating and long-term retrieval components that interfere with habit learning. We speculate about the meaning of this dissociation. Specifically, we suggest that executive control (EF2) competes with the speed of habitual responding, whereas working memory capacity (EF1) supports the accuracy of implicit learning. However, more targeted studies are warranted to empirically validate this theoretical perspective.

Taken together, these findings suggest that while the competitive relationship between specific goal-directed control (EF2) and habit-based processes appears to be a generalizable feature of cognitive functioning—independent of gaming habits or behavioral addiction risk— there are also group-specific interactions involving working memory storage (EF1). In particular, the positive relationship between EF1 and habit learning in the non-gaming and gaming disorder risk groups may reflect compensatory mechanisms that support habit learning when working memory resources are available. A more in-depth analysis—especially at the neural level—could provide valuable insights into the mechanisms of habitual processes and executive functions, offering a more complete understanding of how these interactions play out across different groups and individual profiles.

### 4.4 Limitations

When interpreting our findings, it is important to consider the limitations of this study. Although the IGDT-10 questionnaire, which we used for group classification, is a widely used and reliable assessment tool based on DSM-5 criteria (Király et al., 2019; Yoon et al., 2021), it does not have standalone diagnostic value. Therefore, we cannot definitively determine whether participants met the clinical criteria for gaming disorder; we can only infer that they exhibited a high risk of it. Moreover, as the IGDT-10 is a self-report questionnaire, the lack of self-awareness and the influence of social desirability may distort participants’ responses. In addition, we did not assess the duration of the reported symptoms, which limits our ability to evaluate their clinical significance. Future studies should also consider evaluating the types of video games participants engage with, as different genres have been linked to improvements in distinct cognitive domains (Deleuze et al., 2017; Hong et al., 2022). An important open question remains whether gaming induces changes in cognitive functioning, whether individuals with pre-existing cognitive strengths are more likely to engage in gaming, or whether cognitive deficits are exacerbated in the case of gaming disorder (Waris et al., 2019). Since our study employed a cross-sectional design, longitudinal research is needed to clarify these directional relationships by tracking cognitive changes over time. Finally, a limitation of our study is that we used convenience sampling to recruit participants, which limits the generalizability of our findings. Future research should aim to use more representative sampling methods to improve external validity, ideally with larger sample sizes, as our sample was relatively small.

### 4.5 Conclusions

This was the first study to comprehensively examine the cognitive profiles of individuals at risk of gaming disorder, recreational gamers, and non-gamers, offering new insights into how gaming habits relate to cognitive functioning. Our findings indicate that playing video games regularly in itself is not linked to cognitive difficulties. Instead, it is the presence of problematic, addictive gaming patterns that is associated with cognitive underperformance—particularly in working memory and impulse control—differentiating those at risk of gaming disorder from recreational gamers. In contrast, recreational gamers showed enhanced attentional control and faster responses without accuracy loss, indicating potential cognitive benefits of non-problematic gaming. Notably, we moved beyond examining executive functions in isolation by highlighting the competition between goal-directed and habitual processes. This interaction varied across groups, reflecting how individual gaming habits shape cognitive functioning. These results highlight the importance of not demonizing regular gaming itself but rather focusing on identifying and addressing maladaptive patterns that distinguish recreational gaming from gaming disorder. By mapping these distinct cognitive profiles, our study advances a more nuanced, functional understanding of gaming behavior and lays a foundation for targeted interventions.

## Data availability

All data are available at the following link: https://osf.io/2658w/?view_only=52521253360f423998eb7c0a3b00d978

## CRediT authorship contribution statement

**KB:** Data curation, Formal analysis, Methodology, Project administration, Visualization, Writing – original draft. **ZVP**: Data curation, Formal analysis, Methodology, Project administration, Visualization, Writing – original draft. **TV:** Formal analysis, Software, Supervision, Methodology, Writing – review & editing. **BCF**: Formal analysis, Methodology, Writing – review & editing. **OK:** Conceptualization, Writing – review & editing**. ZD:** Conceptualization, Writing – review & editing**. DN:** Conceptualization, Funding acquisition, Formal analysis Methodology, Resources Writing – review & editing, Supervision. **BK:** Conceptualization, Data curation, Funding acquisition, Investigation, Methodology, Project administration, Resources, Supervision Writing – review & editing.

## Declaration of generative AI use

In the preparation of this manuscript, ChatGPT was used for language and grammar editing to improve clarity and readability. All intellectual content, data analysis, and conclusions are the sole responsibility of the authors.

