## Supplementary Materials for "Game On or Gone Too Far? Executive Functioning and Habit Learning in Problematic vs. Recreational Gamers"

**S1. Quality control of the data**

A total of 21 participants were excluded from the analysis because they did not meet the IGDT cutoff scores for either the recreational or the gaming disorder group. Additionally, 14 participants were excluded from the 1-Back task as statistical outliers, and 3 due to data saving errors. For the 2-Back task, 4 participants were excluded as outliers and 2 due to data saving errors. In the Go/No-Go task, 2 participants were excluded as outliers and 1 due to a data saving error. In the Alternating Serial Reaction Time task, 14 participants were excluded due to data saving errors. One participant was excluded from the Digit Span task and two from the Counting Span task due to data recording issues. Finally, one participant was excluded from the Card Sorting task due to a data saving error.

**S3. How the Alternating Serial Reaction Time task (ASRT) measures implicit pattern learning**

The Alternating Serial Reaction Time (ASRT) task consists of an alternating sequence, where every first stimulus follows a fixed pattern, and every second is randomly selected from four possible locations (e.g., 2 x 4 x 1 x 3 x, with 'x' denoting random stimuli). This structure results in high- and low-probability triplets: high-probability triplets (e.g., 2 x 4 or 4 x 1) occur more frequently as their first and last elements can come from either the fixed pattern or random stimuli. In contrast, low-probability triplets (e.g., 4 x 2 or 1 x 4) only form when both outer elements are random. Consequently, high-probability triplets appear approximately five times more often than low-probability ones.

In terms of occurrence rates, high-probability triplets appear with a 4% probability at each keystroke and account for 62.5% of all triplets within a block, whereas low-probability triplets appear with a 0.8% probability per keystroke, comprising 37.5% of the total. This frequency difference leads to greater habit learning of high-probability triplets. Consequently, if statistical learning functions effectively, participants should exhibit a stronger performance advantage for high-probability triplets compared to low-probability ones.

Our analysis excluded pattern types known as 'trills'—alternating number sequences (e.g., 1-2-1). We also removed direct repetitions (e.g., 2-2-2), as both trills and repetitions may not accurately reflect the cognitive process under investigation. This exclusion accounted for participants' potential preexisting biases toward these patterns, which could skew the data and compromise the integrity of our findings. Additionally, incorrect responses were excluded from the RT analysis.

**S2. Comparison of NG, RG and GDR groups in Age, Childhood SES, Current SES, Gender, Educational Level and Place of Residence**

Table S1.

*Comparison of NG, RG and GDR groups in Age, Childhood SES, Current SES, Gender, Educational Level and Place of Residence*

| Variable | Statistics  H/χ2 | *df* | *p* |
| --- | --- | --- | --- |
| Age | 35.8 | 2 | **< .001** |
| Childhood SES | 2.64 | 2 | .267 |
| Current SES | 0.02 | 2 | .992 |
| Gender | 14.31 | 2 | **< .001** |
| Education | 22.28 | 8 | **.004** |
| Place of residence | 3.431 | 6 | .753 |

**S3. Control variables included in ANCOVA analysis**

In accordance with the assumptions of ANCOVA, we included only those control variables in each analysis that showed a significant association with the given dependent variable. These associations were tested using linear regression. Table S2 presents the relationships between the control variables and the dependent variables, as well as their interactions with group. When none of the control variables showed a significant association with the dependent variable, a one-way ANOVA was conducted instead—using either the parametric or non-parametric version, depending on whether the statistical assumptions were met.

**Table S2.**

*Summary of group-level effects, covariates, and interactions across cognitive variables.*

| Variable | Effect Type | df | Test Statistic | p | Effect Size |
| --- | --- | --- | --- | --- | --- |
| Go/No-Go Hit | Group | 2, 102 | 4.572 | .013* | .060^a^ |
|  | Gender | 1, 102 | 6.233 | .014* | .060 |
|  | Age | 1, 102 | 13.743 | < .001*** | .122 |
|  | Group x Gender | 2, 102 | .222 | .801 | .002 |
|  | Group x Age | 2, 102 | .522 | .595 | .010 |
| Go/No-Go False Alarms | Group | 2 | .8432 | .656 | -.011^b^ |
| Go/No-Go d-prime | Group | 2, 108 | 1.226 | .297 | .022^a^ |
| CST Perseverative Errors | Group | 2, 100 | 2.765 | .068 | .064^a^ |
| CST Perseverative Errors | Education | 4, 100 | 1.959 | .107 | .073 |
| CST Perseverative Errors | Group x Education | 5, 100 | .211 | .957 | .010 |
| DSPAN | Group | 2 | 7.478 | .024* | .050^b^ |
| CSPAN | Group | 2 | 9.953 | .007** | .073^b^ |
| 1-back Hit | Group | 2 | 4.072 | .131 | .022^b^ |
| 1-back False Alarms | Group | 2, 91 | 6.415 | .003** | .125^a^ |
| 1-back False Alarms | Gender | 1, 91 | 4.578 | .035* | .048 |
| 1-back False Alarms | Group x Gender | 2, 91 | .889 | .415 | .019 |
| 1-back d-prime | Group | 2 | 4.072 | .131 | .022^b^ |
| 2-back Hit | Group | 2, 105 | .748 | .476 | .014^a^ |
| 2-back False Alarms | Group | 2, 96 | .407 | .667 | .019^a^ |
| 2-back False Alarms | Education | 4, 96 | 3.016 | .022* | .112 |
| 2-back False Alarms | Group x Education | 4, 96 | .140 | .967 | .006 |
| 2-back d-prime | Group | 2, 96 | .752 | .474 | .026^a^ |
| 2-back d-prime | Education | 4, 96 | 1.849 | .126 | .072 |
| 2-back d-prime | Group x Education | 4, 96 | .306 | .873 | .013 |
| EF1 | Group | 2, 85 | 6.181 | .003** | .089^a^ |
| EF1 | Age | 1, 85 | .515 | .475 | .006 |
| EF1 | Group x Age | 2, 85 | 1.405 | .251 | .032 |
| EF2 | Group | 2 | 3.829 | .147 | .021^b^ |

*Note.* The table summarizes the effects of group, covariates (e.g., age, gender, education), and their interactions on the analyzed cognitive outcomes. Test statistics, degrees of freedom (df), p-values, and effect sizes are reported. Effect sizes are marked as b for eta squared and a for partial eta squared. Significance levels are denoted with asterisks (**p* < .05, ***p* < .01, ****p* < .001). ANCOVA or non-parametric alternatives were used depending on assumption checks for each variable.

**S4. Factor analysis of executive function tasks**

**S4.1 Method of the factor analysis**

We first aimed to determine the potential latent structure of our sets of executive functions (EF) measures in a data-driven manner. Therefore, we investigated the factor structure of the EF measures, using maximum likelihood exploratory factor analysis (ML EFA) with varimax rotation, as implemented in the psych package in R (Revelle, 2022), with default settings. To aid interpretation, the scores that reflect error percentages were subtracted from 100, so that higher values represent better performance in all variables. To assess the factorability of the data, we utilized 3 complementary approaches (Dziuban & Shirkey, 1974). Firstly, we inspected the off-diagonal elements of the anti-image covariance matrix. If the dataset is appropriate for factor analysis, these elements should be all above 0.50 (Dziuban & Shirkey, 1974). Secondly, we computed the Kaiser-Meyer-Olkin (KMO) test of sampling adequacy (Kaiser, 1970). The higher the overall KMO index, the more appropriate a factor analytic model is for the data. The original cut-off recommended by Kaiser (1970) is 0.60, but other authors have also suggested 0.50 (Dziuban & Shirkey, 1974). Finally, we performed Bartlett’s test of sphericity, which tests the hypothesis that the sample correlation matrix came from a multivariate normal population in which the variables of interest are independent (Bartlett, 1950). Rejection of the hypothesis is taken as an indication that the data are appropriate for analysis.

We determined the number of factors to extract using Horn’s parallel analysis (Horn, 1965). This approach is based on comparing the eigenvalues of factors of the observed data with those of random data from a matrix of the same size. Factors with higher eigenvalues in the observed, than in the random data are kept. We used a more stringent criteria, and compared the observed eigenvalues to the 95th percentile, instead of the mean of the simulated distributions. Furthermore, our use of ML EFA also allowed us to calculate multiple fit indices of the applied factor analytic models. Following the recommendations of Fabrigar et al. (1999) and Hu and Bentler (1999), we chose to focus on the Root Mean Square Error of Approximation (RMSEA) and the Standardized Root Mean Squared Residual (SRMR). According to a commonly used guideline, RMSEA values less than 0.05 constitute good fit, values in the 0.05–0.08 range acceptable fit, values in the 0.08–0.10 range marginal fit, and values greater than 0.10 poor fit. Similarly, SRMR values below 0.08 are generally considered indicators of good model fit (Hu & Bentler, 1999). After we determined the number of factors to extract, participant level factor scores were calculated for all factors, based on Thomson’s (1939) regression method.

**Sx.2 Results of the factor analysis**

We established the factorability of the data using 3 approaches. Firstly, the diagonals of the anti-image correlation matrix of the data were all over 0.5, which suggests good factorability. Secondly, the overall Kaiser-Meyer-Olkin (KMO) measure of sampling adequacy was 0.61, which is above commonly used cutoff levels (Dziuban & Shirkey, 1974; Kaiser, 1970). Finally, Bartlett’s test of sphericity was significant (χ2 (15) = 70.307, *p* < .001), suggesting adequate factorability. Based on this, the data were deemed appropriate for factor analysis.

To determine the number of factors to extract, we relied on parallel analysis, and goodness of fit indices. Parallel analysis suggested that two factors were extractable, based on the comparison of the eigenvalues with randomly generated data (Fig. S1). Thus, the two factor solution was selected.

**Fig. S1.**

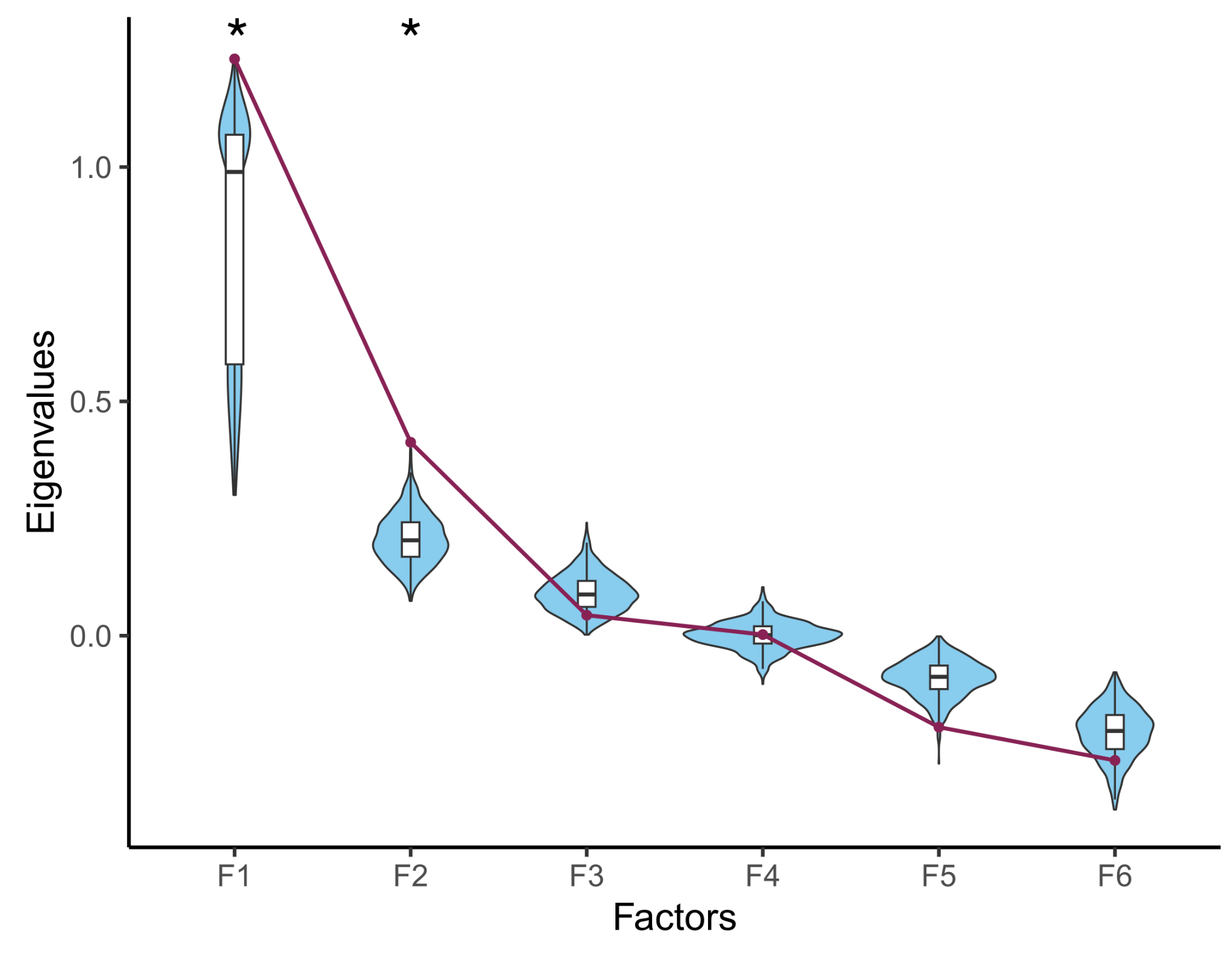
*Parallel analysis of factors.*

The observed eigenvalues are indicated by red dots. These were compared to the distribution of eigenvalues obtained from simulated data, indicated by violin plots and boxplots. Stars indicate factors for which the observed eigenvalue is larger than the 95th percentile of the simulated distributions, and thus were deemed extractable.

These two factors explained 20% and 17% of variance respectively, leading to a total explained variance of 37%. Goodness of fit indices were also indicative of good fit (RMSEA = 0.000, 95% CI = [0.000, 0.0877]; SRMR = 0.024). Factor loadings are presented in Table 5 in the main text of the manuscript. Factor 1 had high loadings from the simple working memory tasks (CSPAN, DSPAN), whereas Factor 2 comprised primarily complex working memory and inhibition (1-back, 2-back, Go/No-Go), confirming the pattern of bivariate correlations (Fig. S2). Surprisingly, WCST seemed relatively independent of the other EF tasks. We calculated factor scores for each subject, which are used in the analyses below.

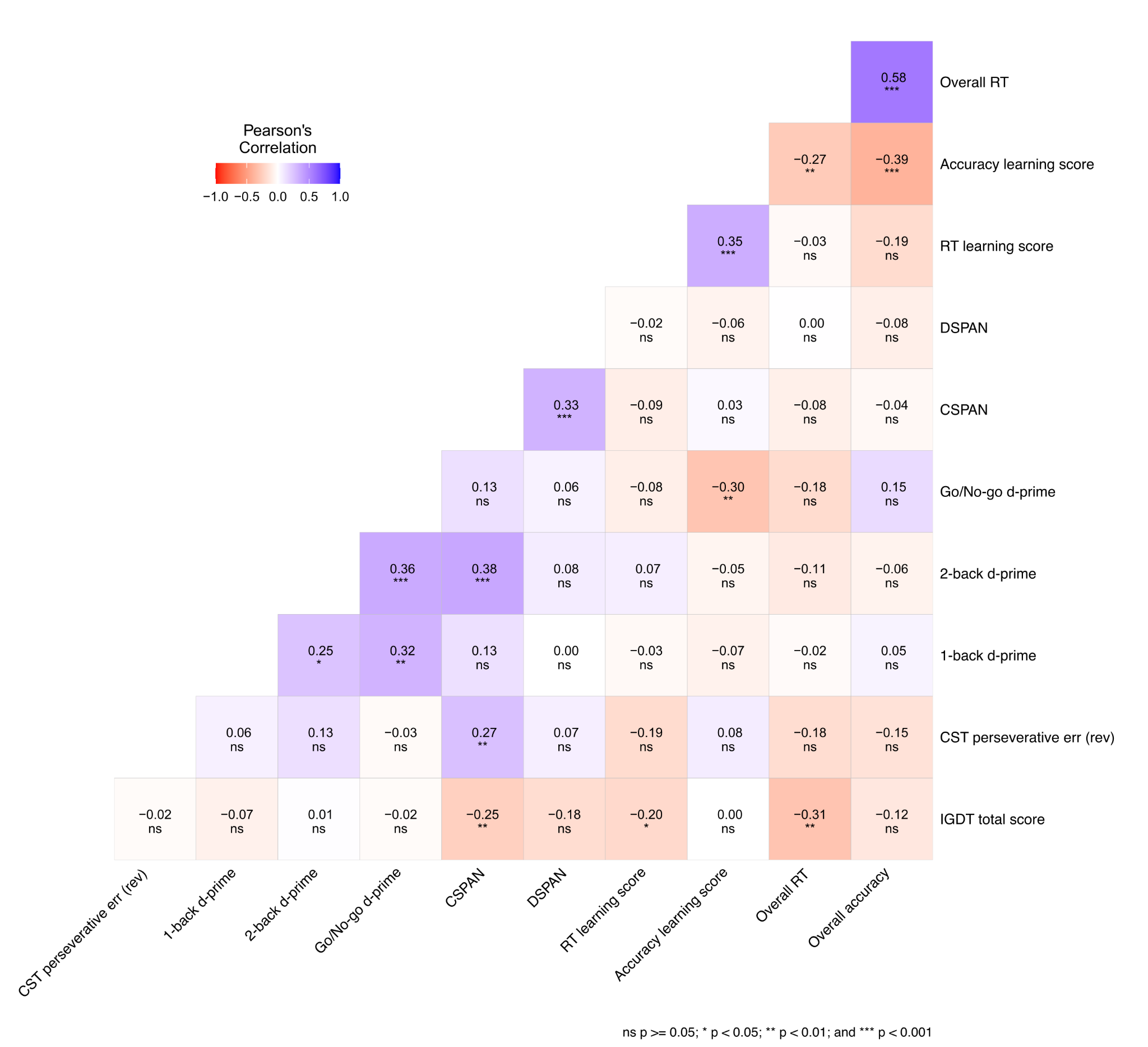
 **Fig. S2. Bivariate Pearson’s correlations between the individual EF measures and average ASRT learning scores.**

More negative and more positive correlations are indicated by red and blue backgrounds, respectively. P values are not corrected for multiple comparisons.

**S5. Habit learning and executive functions in the three groups**

**S5.1 Method of the generalized linear mixed model analyses**

In reporting our generalized linear mixed model (GLMM) analyses, we follow best practice guidelines proposed by Meteyard and Davies (2020). All pre-processing and statistical analyses were performed in R 4.2.1. GLMMs were fit with the mixed function from the afex package (Singmann et al., 2023). No a priori power analysis was done for the study. Our primary independent variables of interest were Epoch, Triplet type, Group, and EF1 and EF2 factor scores. Triplet type was a factor variable, effects coded to reflect the high- and low-probability triplet categories, with low-probability triplets being the reference category. Epoch was a factor variable, coded to reflect Epochs (1 to 4), with the first Epoch being the reference category in all models. EF1 and EF2 factor scores were continuous and centered. Covariates of Age and Education were also continuous and centered, whereas Gender was a factor with Male being the reference category. Following general recommendations, we started with the maximal random effects structure, specifying correlated by-participant intercept and slopes for all within-subjects variables, and simplified in case convergence issues or singular fits. This led to a final model containing correlated by-participant intercept and slopes for Epoch and Triplet type in the case of the RT model, and by-participant uncorrelated intercept and slopes for Epoch in the accuracy model. Thus, our RT model corresponded to the following general form in lmer syntax:

Log(RT) ~ Epoch*Triplet type*(Group*EF1 + Group*EF2 + Education + Age + Gender) + (Epoch + Triplet type | Participant)

Our accuracy model was the same, except with the simpler random effects structure:

Accuracy(0 vs 1) ~ Epoch*Triplet type*(Group*EF1 + Group*EF2 + Education + Age + Gender) + (Epoch || Participant)

Covariates were included if: 1) they shared variance with the outcome (overall RT or accuracy), but not our predictor variables, in order to lower residual variance in the outcome and lead to greater statistical power; 2) if they shared variance with both the outcome and predictor variables, in order to mitigate likely confounding; 3) if they were markedly different between the groups (Table S1; Carlson & Wu, 2012; Wysocki et al., 2022). Assumptions regarding linearity, homoscedasticity and normality of residuals were evaluated by scatterplots and QQ plots of residuals. Assumptions of linearity and homoscedasticity were met in each case, but some of the models presented with non-normal residuals. We note however, that fixed effects estimates of LMMs have been shown to be remarkably robust against violation of this assumption (Schielzeth et al., 2020). Therefore, we deemed our models adequate.

We report sample sizes in terms of total number of data points and of sampling units, random effects estimates, Nakagawa’s marginal and conditional R2 (Nakagawa & Schielzeth, 2013), and the adjusted ICC in summary tables. Post-hoc contrasts were conducted with the emmeans R package, after regridding (Lenth, 2022). Significant interaction effects with continuous variables were interpreted by estimating performance at low (mean-1SD) and high (mean+1SD) levels of the relevant continuous variable. For inference about fixed effects, we used Type III tests (for the RT model) or Likelihood ratio tests (for the accuracy model), relying on comparing nested models with the effect of interest either included or removed. For RT models, degrees of freedom were obtained using the Satterthwaite approximation (Satterthwaite, 1941), for the accuracy models, they are asymptotic, making the test statistic a z statistic, instead of t statistic (Singmann & Kellen, 2019). Figures were created with ggplot2 (Wickham, 2016). An alpha level of .05 was applied to all analyses. P values of all post hoc tests were adjusted with the Šidák method in case of multiple comparisons across Epochs (Šidák, 1967).

**S5.2 Results of the generalized linear mixed model analyses**

We first evaluated habit learning and visuomotor performance in RT, and their relationship with EF factor scores in the three groups of participants (Fig. S3A). To do this, trial-level RT (log transformed) was used as the outcome variables in GLMMs, containing fixed effects of Epoch (Factor: 1, 2, 3, 4), Triplet type (Factor: High, Low), Group (Factor: NG, RG, GDR), and EF1 and EF2 (continuous and centered), along with their higher order interactions, with participant-specific correlated intercepts and slopes for the main effects of Epoch and Triplet type. Our selected control variables of Age (continuous and centered), Gender (Factor: Male, Female) and Education (continuous and centered) were also included as fixed effects in interaction with the manipulated independent variables of Epoch and Triplet type (see Methods for more details). Throughout the presentation of the results, main effects of Epoch are interpreted as indicating visuomotor performance, independent of statistical regularities, while main effects and interactions of Triplet type are interpreted as indicating habit learning and its trajectory during the task. Henceforth, the term learning score always denotes performance differences between High and Low triplets, obtained from contrasts, with positive sign always corresponding with better learning. Post hoc tests are done using model-estimated marginal means (Lenth, 2022). Note that EF1 and EF2 are incorporated into the model as continuous variables, and Low and High EF subgroups are created only for post hoc testing and visualization.

In this model, there was as statistically significant main effect of Epoch (F_(3,68.89)_ = 75.55, p < .001), indicating highest visuomotor performance in the fourth epoch (epoch 1: 376 ms, 95% CI = [367, 385]; epoch 2: 374 ms, 95% CI = [365, 384]; epoch 3: 373 ms, 95% CI = [364, 382]; epoch 4: 350 ms, 95% CI = [341, 359]; epoch 4 significantly lower than all preceding epochs, all ps < .001; no other pairwise comparison significant, all ps > .420). Given the lack of a significant main effect of Group (F_(2,68.12)_ = 2.27, p = .111), or an Epoch × Group interaction (F_(6,68.97)_ = 1.56, p = .171), overall visuomotor performance did not differ between groups. The main effect of Triplet type (F_(1,67.16)_ = 70.35, p < .001) and the Epoch × Triplet type interaction (F_(3,91240.02)_ = 12.72, p < .001) were both statistically significant, indicating increasing habit learning with time (learning scores in epoch 1: 1.82 ms, 95% CI = [-0.79, 4.44]; epoch 2: 7.09 ms, 95% CI = [4.47, 9.70]; epoch 3: 8.91 ms, 95% CI = [6.34, 11.49]; epoch 4: 10.26 ms, 95% CI = [7.87, 12.64]; epoch 1 significantly lower than all other epochs, all ps < .009; no other pairwise comparison significant, all ps > .230).

The model also resulted in a statistically significant Epoch × Triplet type × Group × EF2 interaction (F_(6,58994.06)_ = 2.44, p = .023) (Fig. S3B). This stemmed from a different trajectory of habit learning in the RG group depending on EF2 scores, whereas in the other two groups EF2 had no association with learning scores (Fig. S2). In the RG group, learning scores were significantly higher in epoch 3 in individuals with lower EF2 (estimate of learning score difference between low and high EF2: 12.59 ms, 95% CI = [3.18, 21.99], p = .001), with no difference in any other epoch (all ps > .148). In both the NG and GDR groups, there were no significant differences between lower and higher EF2 individuals in any epoch (all ps > .163).

No further main effect or interaction involving either Group or EF factor scores was statistically significant. Overall, these results suggest that visuomotor performance was comparable between the groups, and that there was stronger competition between EF2 and habit learning in the RG group than in either the NG or the GDR groups.

*Accuracy*

Next, we investigated habit learning and visuomotor performance in accuracy (0: incorrect vs 1: correct), using a binomial GLMM (Fig. S4). The fixed effects for the accuracy model were exactly the same as in the RT model, however the random effects structure needed to be simplified to contain uncorrelated by-participant random intercepts and slopes for Epoch only.

In this model, there was a statistically significant main effect of Epoch (*χ^2^* (3) = 15.09, p = .002), which indicated significantly lower accuracy in epoch 3 compared to epoch 1 and epoch 4 (epoch 1: 94.7 %, 95% CI = [93.6, 95.6]; epoch 2: 94.4 %, 95% CI = [93.2, 95.3]; epoch 3: 94.2 %, 95% CI = [93.0, 95.2]; epoch 4: 94.9 %, 95% CI = [93.8, 95.8]; epoch 4 significantly higher than epoch 3, p_uncorrected_ = .019 (p_Sidak_ = .108); no other pairwise comparison significant, all ps > .072). The main effect of Triplet type was not statistically significant (*χ^2^* (1) = 1.94, p = .164), but the Epoch × Triplet type interaction was (*χ^2^* (3) = 8.22, p = .042). This stemmed from increased habit learning with time (learning scores in epoch 1: 0.74 %, 95% CI = [0.01, 1.48]; epoch 2: 1.43 %, 95% CI = [0.64, 2.22]; epoch 3: 1.66 %, 95% CI = [0.86, 2.46]; epoch 4: 2.03 %, 95% CI = [1.24, 2.81]; epoch 1 significantly lower than epoch 4, p_uncorrected_ = .016 (p_Sidak_ = .094); no other pairwise comparison significant, all ps > .091).

We next interpret the complex associations between executive function factor scores and task performance (Fig. S4). Regarding EF1, we observed statistically significant Triplet type × EF1 (*χ^2^* (1) = 8.93, p = .003) and Triplet type × Group × EF1 (*χ^2^* (2) = 6.18, p = .045) interactions. These stemmed from significant learning scores only in individuals with higher EF1 in the NG (learning scores in low EF1: 0.74 %, 95% CI = [-0.15, 1.64]; high EF1: 1.97 %, 95% CI = [1.01, 2.87]) and GDR groups (learning scores in low EF1: 0.56 %, 95% CI = [-0.07, 1.18]; high EF1: 2.55 %, 95% CI = [0.018, 4.91]), whereas there was no association between learning scores and EF1 scores in the RG group (learning scores in low EF1: 1.51 %, 95% CI = [0.57, 2.45]; high EF1: 1.80 %, 95% CI = [0.86, 2.73]).

Regarding EF2, there was a significant Triplet type × EF2 interaction (*χ^2^* (1) = 5.55, p = .018), which stemmed from lower learning scores in high EF2 individuals (estimate of learning score difference between low and high EF2: 1.67 %, 95% CI = [0.53, 2.82], p = .004). There were also significant Group × EF2 (*χ^2^* (2) = 7.52, p = .023), Epoch x EF2 (*χ^2^* (3) = 9.98, p = .019) interactions, and some evidence for an Epoch × Group × EF2 (*χ^2^* (6) = 12.04, p = .061) interaction that did not reach our alpha level. These stemmed from generally higher accuracy in individuals with higher EF2 scores in the GDR group, especially in epochs 1 and 2 (estimate of difference between low and high EF2 in epoch 1: 4.83 %, 95% CI = [0.11, 9.54]; epoch 2: 5.22 %, 95% CI = [0.31, 10.12]; epoch 3: 4.52 %, 95% CI = [-0.57, 9.62]; epoch 4: 4.59 %, 95% CI = [0.07, 9.25]), whereas there was a weaker and less consistent relationship in the RG group (estimate of difference in general between low and high EF2 in epoch 1: 3.92 %, 95% CI = [-1.06, 8.91]; epoch 2: 4.09 %, 95% CI = [-1.09, 9.28]; epoch 3: 4.11 %, 95% CI = [-1.22, 9.43]; epoch 4: 3.52 %, 95% CI = [-1.68, 8.72]) and the NG group (estimate of difference between low and high EF2 in epoch 1: -1.04 %, 95% CI = [-4.36, 2.29]; epoch 2: -0.55 %, 95% CI = [-4.35, 3.26]; epoch 3: 0.51 %, 95% CI = [-3.26, 4.27]; epoch 4: 1.27 %, 95% CI = [-2.28, 4.82]).

To summarize, EF1 scores had no relationship with overall accuracy. Instead, they showed a positive association with habit learning ability, but only in the NG and GDR groups, and no association in the RG group. EF2 scores were associated with better overall accuracy in the GDR group only, and generally had a negative association with habit learning in all groups.

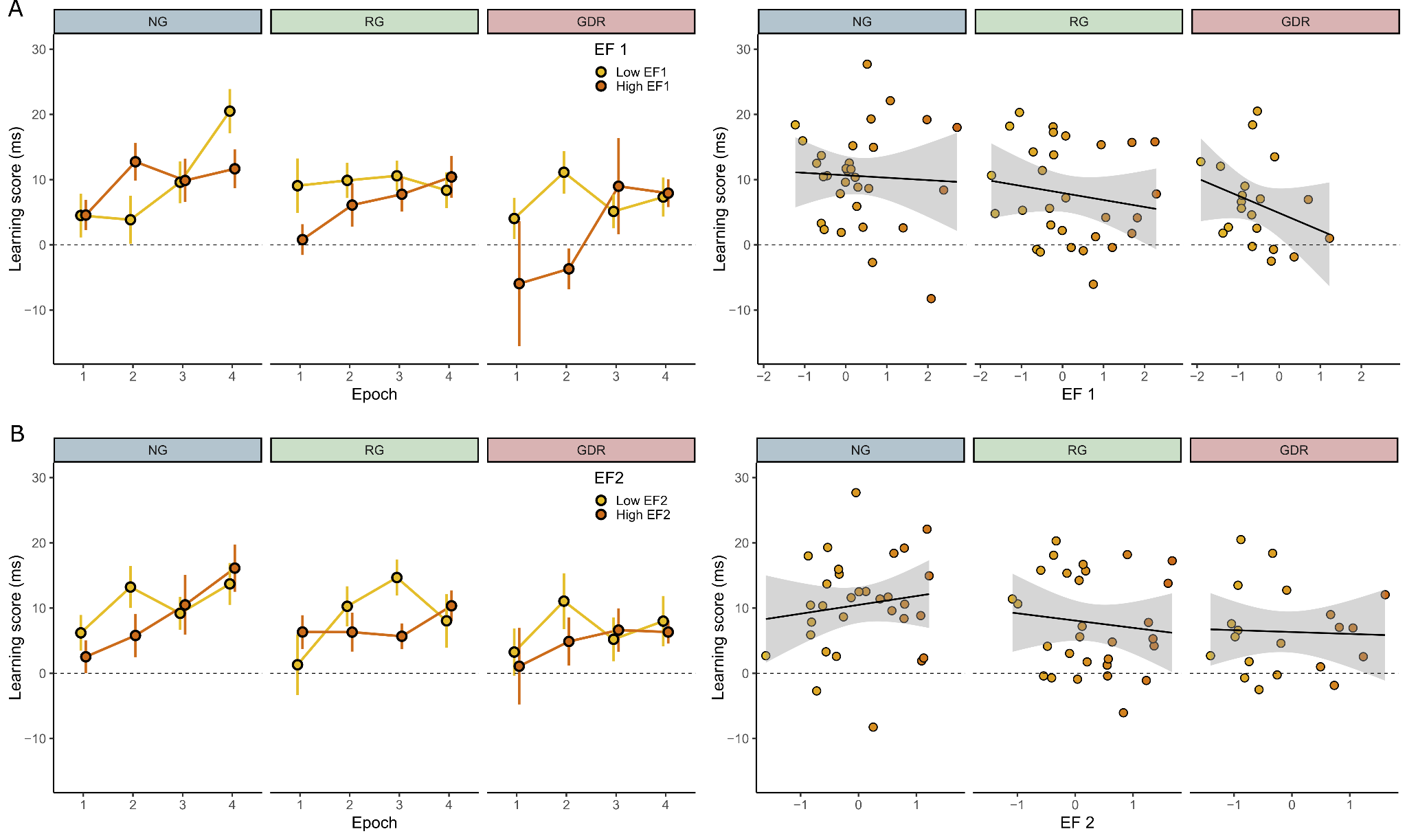

**Fig. S3. Trajectories habit learning scores in the three groups as a function of executive function scores in RT.** **A)** Learning scores as a function of EF1 scores in the three groups. **B)** Learning scores as a function EF2 scores in the three groups. Figures on the left show the trajectories of learning scores across epochs, points indicate means, error bars indicate 1 SEM, and participants are median split into Low (blue) and High (red) EF groups. Figures on the right are scatterplots showing the relationship between EF scores and average learning scores. Thick black line indicates the line of best fit, with shaded region showing 95% confidence intervals. Each point is one participant, coloring of points indicates EF scores from low (blue hue) to high (red hue).

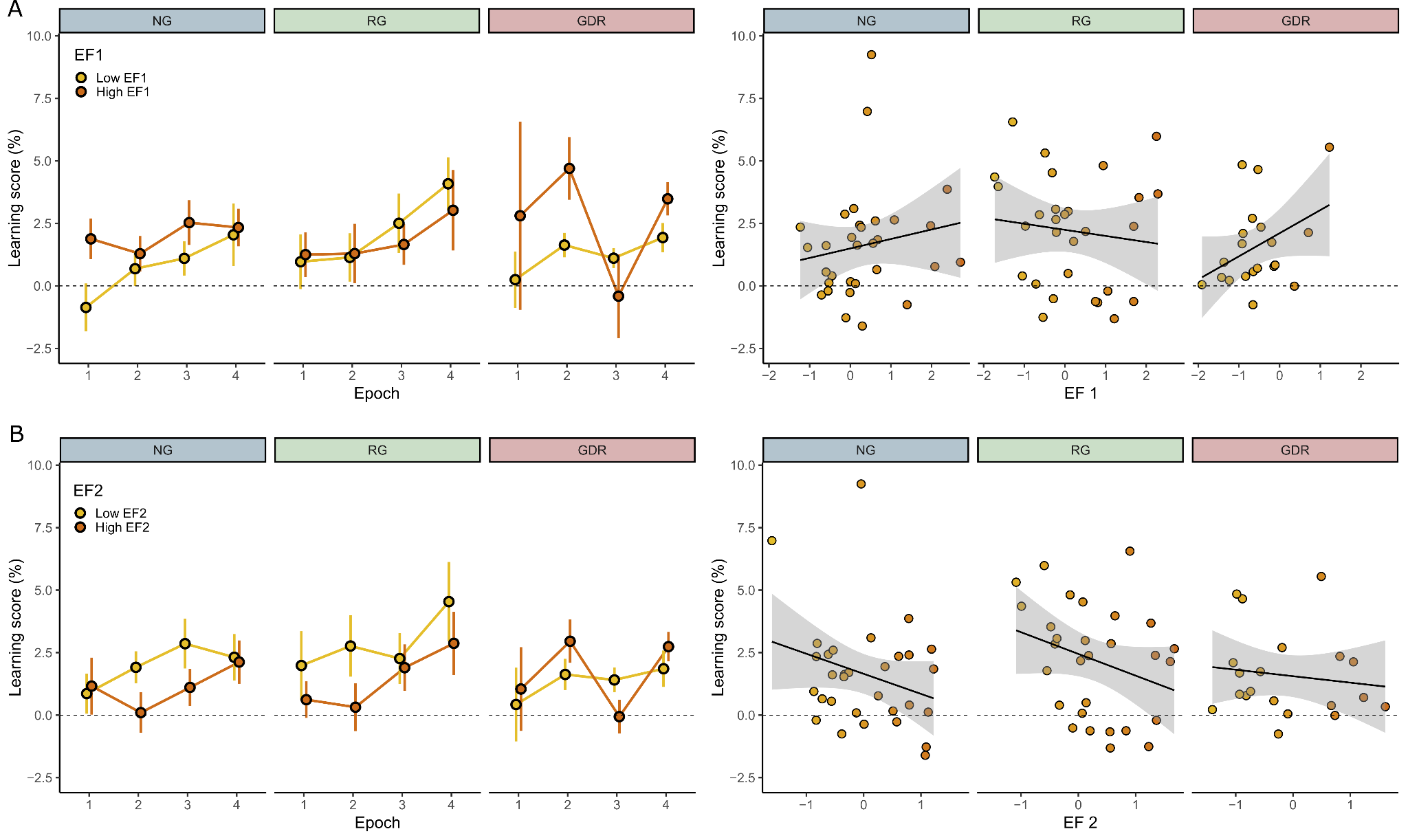

**Fig. S4. Trajectories of habit learning scores in the three groups as a function of executive function scores in accuracy.** **A)** Learning scores as a function of EF1 scores in the three groups. **B)** Learning scores as a function EF2 scores in the three groups. Figures on the left show the trajectories of learning scores across epochs, points indicate means, error bars indicate 1 SEM, and participants are median split into Low (blue) and High (red) EF groups. Figures on the right are scatterplots showing the relationship between EF scores and average learning scores. Thick black line indicates the line of best fit, with shaded region showing 95% confidence intervals. Each point is one participant, coloring of points indicates EF scores from low (blue hue) to high (red hue).
